# The Interplay between Maternal Smoking and Genes in Offspring Birth Weight

**DOI:** 10.1101/2020.10.30.20222844

**Authors:** Rita Dias Pereira, Cornelius A. Rietveld, Hans van Kippersluis

## Abstract

It is well-established that both the child’s genetic endowments as well as maternal smoking during pregnancy impact offspring birth weight. In this paper we move beyond the nature *versus* nurture debate by investigating the interaction between genetic endowments and this critical prenatal environmental exposure – maternal smoking – in determining birth weight. We draw on longitudinal data from the Avon Longitudinal Study of Parents and Children (ALSPAC) study and replicate our results using data from the UK Biobank. Genetic endowments of the children are proxied with a polygenic score that is constructed based on the results of the most recent genome-wide association study of birth weight. We instrument the maternal decision to smoke during pregnancy with a genetic variant (rs1051730) located in the nicotine receptor gene CHRNA3. This genetic variant is associated with the number of cigarettes consumed daily, and we present evidence that this is plausibly the only channel through which the maternal genetic variant affects the child’s birth weight. Additionally, we deal with the misreporting of maternal smoking by using measures of cotinine, a biomarker of nicotine, collected from the mother’s urine during their pregnancy. We confirm earlier findings that genetic endowments as well as maternal smoking during pregnancy significantly affects the child’s birth weight. However, we do not find evidence of meaningful interactions between genetic endowments and an adverse fetal environment, suggesting that the child’s genetic predisposition cannot cushion the damaging effects of maternal smoking.

## 1. Introduction

Early life experiences matter: The fetal origins literature has convincingly established that a wide-range of prenatal exposures including stress, pollution, maternal risky health behaviors, and nutrition have long-term consequences (Currie, 2009; Almond and Currie, 2011; Almond et al., 2018). Likewise, the behavior genetics literature has established that all relevant life outcomes are partially heritable, with the vast majority of outcomes being influenced by many genetic variants with each a tiny effect (Turkheimer, 2000; Chabris et al., 2015; Polderman et al., 2015). The current consensus is therefore that both genes and the environment matter, and that the traditional nature *versus* nurture debate is obsolete (Turkheimer, 2000; Heckman, 2007). However, whereas it is repeatedly argued that life outcomes result from a complex interplay between genes and environment (Rutter, 2006; Almond et al., 2018), estimations of the actual interaction between genes and the environment remain rare.

In this paper we move beyond the nature versus nurture debate by investigating how birth weight is influenced by the interaction between genes and a critical prenatal environmental exposure: maternal smoking. Birth weight is an important predictor of newborn and infant survival, and is associated with later life health and outcomes such as cognitive development, educational attainment, and earnings (Black et al., 2007; Royer, 2009; Figlio et al., 2014; Bharadwaj et al., 2018; Trejo, 2020). As a consequence, it also affects health and socio-economic outcomes of the next generation (Currie and Moretti, 2007). Birth weight is the most commonly used barometer among pregnancy outcomes (Conti et al., 2018), and it comes with the advantage (compared to later-life outcomes) that its value is easily attributed to genes and environmental exposures in a specific time period, i.e., the 9-month period in-utero.

Previous studies have convincingly established that genetic endowments influence the risk of being born with low birth weight (Horikoshi et al., 2013, 2016; Warrington et al., 2019). Likewise, a body of literature points at maternal smoking during pregnancy as a significant and important determinant of birth weight (Sexton and Hebel, 1984; Kramer, 1987; Hamilton, 2001; Ringel and Evans, 2001;Bharadwaj et al., 2014; Lien and Evans, 2005; Banderali et al., 2015; Simon, 2016; Yang et al., 2019).^2^Our innovation compared with this literature is to study the *interaction* between genetic endowments and maternal smoking during pregnancy in determining the child’s birth weight. This represents not merely an advance in the fundamental understanding of how nature and nurture interact, but additionally reveals whether environmental exposures during pregnancy can exacerbate or compensate for genetic disadvantages in determining life outcomes.^3^

Our main analysis sample comes from the Avon Longitudinal Study of Parents and Children (ALSPAC), a UK cohort study of children born in the early nineties. The data are uniquely suited to answer our research question since both the mothers and their children are genotyped, a nurse administered the child’s birth weight, and maternal smoking was not just self-reported but additionally biochemically validated using biomarkers. The main limitation of the ALSPAC data is that the sample is relatively small – around 5,000 mother-child pairs for whom we observe both genotypes, and only around 2,500 mother-child pairs for whom the nicotine biomarker is available. Therefore, we additionally exploit data from the much larger UK Biobank. In this sample, we have to rely on some proxy- and self-reported variables, but its size (around 250,000 individuals) allows us to replicate our key findings.

To proxy genetic endowments, we construct a birth weight polygenic score using the results of the latest genome-wide association study (GWAS) on birth weight (Warrington et al., 2019). This polygenic score represents the best linear genetic predictor for birth weight, and can be interpreted as one’s genetic predisposition for high or low birth weight. In empirically establishing how the effect of maternal smoking differs by the child’s genetic endowments, we have to address two commonly discussed issues with regards to smoking. First, relying on self-reported data about smoking is problematic given the stigma associated with this act. Especially during a pregnancy, it is likely that women misreport their smoking habits in surveys. In order to tackle this issue we compare the self-reported measures of smoking with a measure of cotinine, a biomarker of nicotine, collected from the mothers while they were pregnant. This approach has been used before to determine actual smoking during pregnancy (Tappin et al., 1997; Lindqvist et al., 2002) and to study the impact of smoking on newborns’ health (Li et al., 1993; Wang et al., 1997). In our data, the prevalence of self-reported smoking is 22% while 31% of the mothers had enough cotinine in their urine during their pregnancy to be considered active smokers. We show that in our data misreporting leads to overestimation of the effect of smoking, which suggests that mothers who smoke and report smoking are different from the ones who smoke but do not report smoking.

Second, smoking mothers are likely to be different from the non-smoking mothers in many ways we cannot account and correct for in a regular regression framework (e.g., Bradford, 2003). Possible factors include how careful they are during their pregnancy, if their pregnancy was planned, and how acquainted they are with the available prenatal care. All these factors are typically unobserved in survey data and hence their omission from the regression model will bias the estimation of the effect of maternal smoking on children’s outcomes. Some studies tackle this issue by exploiting variation in cigarette taxes across states (Evans and Ringel, 1999), the introduction of smoking bans (Bharadwaj et al., 2014) or randomized nurse visits (Mejdoubi et al., 2014) as exogenous factors impacting smoking behavior during pregnancy. In this study, we instrument the number of cigarettes smoked during pregnancy with a genetic instrumental variable (IV): the genetic variant rs1051730 located in the nicotine receptor gene CHRNA3 (Tyrrell et al., 2012; Wehby et al., 2011; Van Kippersluis and Rietveld, 2018a; Yang et al., 2019). This so-called single nucleotide polymorphims (‘SNP’, pronounced ‘snip’) has consistently been associated with the number of cigarettes smoked per day, also during pregnancy (Bierut, 2010; Furberg et al., 2010; Liu et al., 2010) and its biological function is well understood. We find that this SNP consistently reduces offspring birth weight among mothers who smoke, but that it is unrelated to the child’s birth weight among non-smoking mothers. This provides compelling support that the *intensity* of smoking is the single channel through which the IV affects the outcome and thus that this SNP is a suitable instrument for dealing with the endogenous nature of smoking.

Our IV analyses show that, on average, an extra cigarette per day during pregnancy reduces birth weight by 20 to 40 grams. These point estimates are considerably larger than the reduction of 9 to 14 grams we find in the OLS regressions. While our IV estimates could be biased upwards by the underreporting of smoking in the first stage (the effect of SNP rs1051730 on smoking), auxiliary analyses suggest that even under extreme underreporting the point estimates in the IV regression are larger than in the OLS regression. These estimates corroborate earlier findings that maternal smoking during pregnancy reduces birth weight, and go beyond earlier findings in carefully assessing the sign and magnitude of the bias due to the misreporting of smoking.

Despite the robust negative relationship between maternal smoking and birth weight, we do not find evidence of significant heterogeneity in the effect of maternal smoking on birth weight by the child’s genetic predisposition for birth weight. While in ALSPAC the confidence interval of the interaction term does not rule out sizable heterogeneity, our replication analysis in the UK Biobank provides a rather precisely estimated zero for the interaction effect. Our results therefore suggest that maternal smoking as well as genetic endowments influence birth weight in an additive fashion only, and that there is no meaningful moderation effect of one on the other. In other words, even a strong genetic predisposition for high birth weight cannot protect against the negative effects that maternal smoking during pregnancy bring about.

Our study contributes to at least two streams of literature. First, it contributes to the literature on the effects of maternal smoking on offspring birth weight (e.g., England et al., 2001; Tappin et al., 2015). Low birth weight is strongly associated with worse later-life health and socio-economic outcomes, and maternal smoking has been identified as the most significant modifiable risk factor for the incidence of low birth weight in developed countries (Kramer, 1987; Almond et al., 2005). We contribute to this literature by exploiting the unique features of the ALSPAC data that enable us to simultaneously address the underreporting and endogeneity of maternal smoking. In doing so, we obtain a more reliable estimate of the average reduction in birth weight as a result of maternal smoking. Moreover, we are the first to investigate treatment effect heterogeneity related to the child’s genetic predisposition for low birth weight.^4^ Heterogeneity of treatment effects by genotype is not merely important to better understand differential responses to maternal smoking, but additionally has important intergenerational implications. If the effect of maternal smoking consistently affects certain groups with a specific genetic predisposition more, then these effects will propagate through generations, potentially increasing inequalities in future generations.

A second literature to which we contribute is a still small but emerging literature analyzing Gene-by-Environment (*G*×*E*) interactions by exploiting exogenous environmental exposures. Whereas *G*×*E* studies have been around for a long time in the medical and social sciences, estimating meaningful interactions is often prohibited since genes and the environment tend to be correlated. For example, individuals with certain genetic predisposition may self-select into certain environments, and parental genes are not just transmitted to children but additionally shape the rearing environment for their children (Kong et al., 2018). These sources of so-called gene-environment correlation pose a threat to the identification of *G* × *E*, and can only be tackled using exogenous variation in environmental exposures. Here we instrument the environmental exposure maternal smoking by a maternal genetic variant, and we show that this genetic variant is independent of the child’s genetic predisposition for birth weight. As such, by exploiting exogenous variation in the environmental exposure, we contribute to the literature seeking to improve our understanding of the interplay between genes and the environment in shaping life outcomes (e.g. Schmitz and Conley, 2017; Barcellos et al., 2018).

The remainder of our study is organized as follows. The next section describes our main dataset, ALSPAC, and the variables we utilize from it. In section three, we detail our identification strategy. In the fourth section we present our main empirical results, including a number of sensitivity analyses. Section five present the results of our replication study in the UK Biobank. The final section discusses our findings and concludes.

## 2. Data

In this section, we introduce our main dataset and we define and operationalize the variables used in our analyses.

### 2.1. The Avon Longitudinal Study of Parents and Children

The Avon Longitudinal Study of Parents and Children (ALSPAC) is a prospective and longitudinal study of children and parents (Boyd et al., 2013; Fraser et al., 2013). The data collection started during pregnancy with the aim to monitor children from fetal life, through infancy into adolescence and young adulthood. All mothers residing in Bristol, Avon, United Kingdom, with an expected delivery date between April 1, 1991, and December 31, 1992, were eligible to take part in ALSPAC. The mothers and their partners were recruited for the study soon after the confirmation of the pregnancy. Details of their social background, attitudes towards health care, and psychological well-being were obtained by self-completion questionnaires. The clinical course of pregnancy and childbirth were recorded from medical case notes. 14,541 eligible pregnant women were enrolled at baseline. From these pregnancies, 13,988 children were alive at 12 months of age. Maternal blood and urine were collected during pregnancy, and the same samples were later on collected from their children. Finally, both mothers and their children were genotyped. The data set comprises 9,115 genotyped mothers and 9,048 genotyped children. More details about this study can be found in Fraser et al. (2013).^5^

Our analysis of the average treatment effect of maternal smoking is restricted to a baseline sample of 7,598 mother-child pairs, which corresponds to pairs with information about the maternal genotypes as well as about the outcome and main explanatory variable (number of cigarettes smoked per day). We call this baseline sample 1. To study the heterogeneous effects of maternal smoking we further restrict baseline sample 1 to mother-child pairs with non-missing child genotypes, which leaves us with 5,006 mother-child pairs. This constitutes baseline sample 2. Additional analyses further restrict the sample to mother-child pairs for which the mother supplied a urine sample. This sample is necessary to determine the level of cotinine in urine.

### 2.2. Variables

#### Outcome variable

The main outcome of interest in our study is birth weight of the child. The weight was obtained from routine hospital birth records. We restricted our analyses to children alive at 1 year of age, which corresponds in our sample to the exclusion of babies with a birth weight of less than 640g. Table 1 shows that for the 7,598 children in our first baseline sample the mean birth weight is 3,406 grams.

**Table 1.** Descriptive statistics of the analysis sample.

|  | Mean | S.D. | Min. | Max. | N |
| --- | --- | --- | --- | --- | --- |
| **Outcome variable** |  |  |  |  |  |
| Birth weight (grams) | 3,406.24 | 556.60 | 703 | 5,600 | 7,598 |
| **Main explanatory variables** |  |  |  |  |  |
| Smoking regularly | 0.22 | 0.42 | 0 | 1 | 7,417 |
| Smoking regularly (adjusted) | 0.31 | 0.46 | 0 | 1 | 2,777 |
| Number of cigarettes smoked per day | 2.02 | 5.09 | 0 | 51 | 7,598 |
| Number of cigarettes smoked per day (adjusted) | 4.96 | 6.64 | 0 | 51 | 3,784 |
| Cotinine (ng/ml) | 743.76 | 1,923.57 | 0 | 24,674 | 2,844 |
| Birthweight PGS (child) | 0.00 | 1.00 | -3 | 3 | 5006 |
| **Instrumental variable** |  |  |  |  |  |
| rs1051730 | 0.67 | 0.67 | 0 | 2 | 7,598 |
| **Control variables** |  |  |  |  |  |
| Mother's birth weight (grams) | 3,269.01 | 601.43 | 909 | 6,108 | 4,541 |
| Mother's age (years) | 28.47 | 4.66 | 15 | 44 | 7,598 |
| Mother's marital status (Married=1) | 0.78 | 0.41 | 0 | 1 | 7,540 |
| Mother's education (categories) | 3.65 | 1.61 | 0 | 6 | 5,484 |
| Mother's social class (categories) | 2.85 | 1.07 | 1 | 6 | 6,141 |
| Grandmother's education (categories) | 2.45 | 1.29 | 1 | 5 | 4,267 |

S.D.=Standard deviation; Min.=Minimum; Max.=Maximum.

#### Main explanatory variables

The main environmental explanatory variables are measures for maternal smoking. We use a binary indicator for regular smoking during pregnancy as well a continuous variable reflecting the intensity of smoking in terms of the average number of cigarettes smoked per day during pregnancy. Given that our proposed instrumental variable is more closely associated with the number of cigarettes per day rather than the binary indicator of smoking (see section 3 below), most of our analyses will be based on the intensive margin (i.e. number of cigarettes smoked). Both variables are self reported. However, we correct for self-reporting by considering the levels of cotinine in urine collected from the women when they were pregnant.

##### Smoking regularly

At 18 weeks of gestation women were asked to report whether they smoked regularly or not during the first 3 months of their pregnancy. Table 1 shows that 22% of mothers classified themselves as regular smokers.

##### Number of cigarettes smoked per day

During their pregnancy, women were asked to report the number of cigarettes per day they smoke presently. The timing varied from as early as 8 until 42 weeks of gestation. Table 1 shows that the average number of cigarettes smoked per day is 2.02. This number reflects both smokers and non-smokers. The average number of cigarettes smoked per day among smokers is 10.80 (not shown).

##### Cotinine

During the first 3 months of pregnancy, a urine sample was collected and tested for cotinine. Cotinine is the predominant metabolite of nicotine (Langone et al., 1973). It is used as a biomarker for exposure to tobacco smoke.^6^ However, there is discussion about the most appropriate threshold for identifying active smokers from cotinine data. We choose to re-classify women with high cotinine levels during pregnancy as active smokers using a reasonable yet conservative threshold of 100 ng/ml. In Appendix A.1 we provide an in-depth discussion about cotinine thresholds.

##### Smoking regularly (adjusted)

This variable is created by recoding the *Smoking regularly* measure. Mothers are reclassified as smokers if they have more than 100 ng/ml of cotinine in their urine. Compared to the self-reported measure, we classify 31% instead of 22% of the mothers as smokers (see Table 1).^7^

##### Number of cigarettes smoked per day (adjusted)

This variable is created by (i) assuming that people who reported a positive number of cigarettes during pregnancy reported their smoking behavior truthfully (this assumption is relaxed in section 4); (ii) running a simple OLS regression between the number of cigarettes and the value of cotinine in urine for those who reported to have smoked a positive number of cigarettes; and (iii) predicting the number of cigarettes based on that regression for the ones who claimed to have smoked 0 cigarettes but had a cotinine value higher than 100 ng/ml. According to this adjustment, the mean number of cigarettes smoked per day is 4.96 instead of 2.02 (Table 1). Again, this number reflects the number of cigarettes smoked by smokers and non-smokers. The average adjusted number of cigarettes smoked per day among smokers is 10.55.

The other main explanatory variable of interest – the variable that we use to interact with the maternal smoking variables – is the child’s polygenic score (PGS) for birth weight. Recent advances in genetics such as the completion of the Human Genome Project in the early 2000s and the advent of inexpensive genotyping chips have made it possible to identify linkages between a person’s genetic endowments and important life outcomes such as health and socio-economic status (Beauchamp et al., 2011; Visscher et al., 2017). Due to these advances it is now possible to construct credible measures of “nature” for many traits; those measures are called polygenic scores.

##### Birth weight polygenic score

The human genome consists of more than 3.2 billion nucleotides located on 23 pairs of chromosomes (Lehrer and Ding, 2017). These nucleotides come in four varieties: adenine (A), guanine (G), cytosine (C) and thymine (T). Approximately 99.6 percent of the nucleotides are identical between two randomly selected individuals (Kidd et al., 2008). However, there are particular positions in the genome where different nucleotides can be present. The most common type of such genetic variation is called a single nucleotide polymorphism (SNP). Two different nucleotides can be present at a SNP location, and SNPs constitute the main source of genetic differences between individuals. It is common to measure SNPs by counting the number of minor alleles (the nucleotide that occurs least frequently in the population). Hence, a SNP can take the values 0, 1 or 2. There are approximately 85 million SNPs in the human genome with a minor allele frequency >1% (The 1000 Genomes Project Consortium, 2015). In some rare cases, a difference at a specific locus on a chromosome can single-handedly lead to a disease: Huntington’s disease is an example. However, the vast majority of human (behavioral) traits are polygenic, meaning they are influenced by multiple genetic polymorphisms with each a tiny effect (Chabris et al., 2015). A polygenic score is constructed by adding up the individual SNPs, where each SNP is weighted by the strength of the association between the SNP and the outcome variables as estimated in a genome-wide association study (GWAS) (Dudbridge, 2013). The underlying rationale is that based on GWAS results, you can assign weights of relative importance to each SNP. Then, with a polygenic score, one can exploit the joint predictive power of multiple SNPs for a particular outcome.

The predictive power of PGSs increases with the sample size of the underlying GWAS (Dudbridge, 2013). Therefore, meta-analysis techniques are used to make GWASs as large as possible (Visscher et al., 2017). However, to avoid overfitting in PGS analyses, it is necessary to exclude the prediction sample from the GWAS meta-analysis used to construct the PGS. Therefore, in our study, we used the summary statistics of Warrington et al. (2019) to retrieve the relevant SNPs as well as their weights. Importantly, ALSPAC was excluded from this meta-analysis of birth weight. The weights were subsequently corrected for linkage disequilibrium (structural correlation across SNPs in the genome) using the software LDPred (Vilhjálmsson et al., 2015). The resulting PGS is a continuous measure that represents the propensity to be born with a high birth weight as determined by the child’s genotypes. This measure was standardized in such a way that the mean is zero and the standard deviation is 1 in the analysis sample. More details about the genotyping procedure and construction of the PGS can be found in Appendix A.2 and Appendix A.3.

Figure 1 shows that, despite a lot of individual variation, the PGS for birth weight predicts actual birth weight in our sample. This figure also suggests that a linear relationship between the variables provides a reasonable fit with the data. The difference between the bottom and top decile of the PGS is around 400 grams in birth weight, and we can explain around 5% of the variation in birth weight with the PGS.

**Fig 1.**
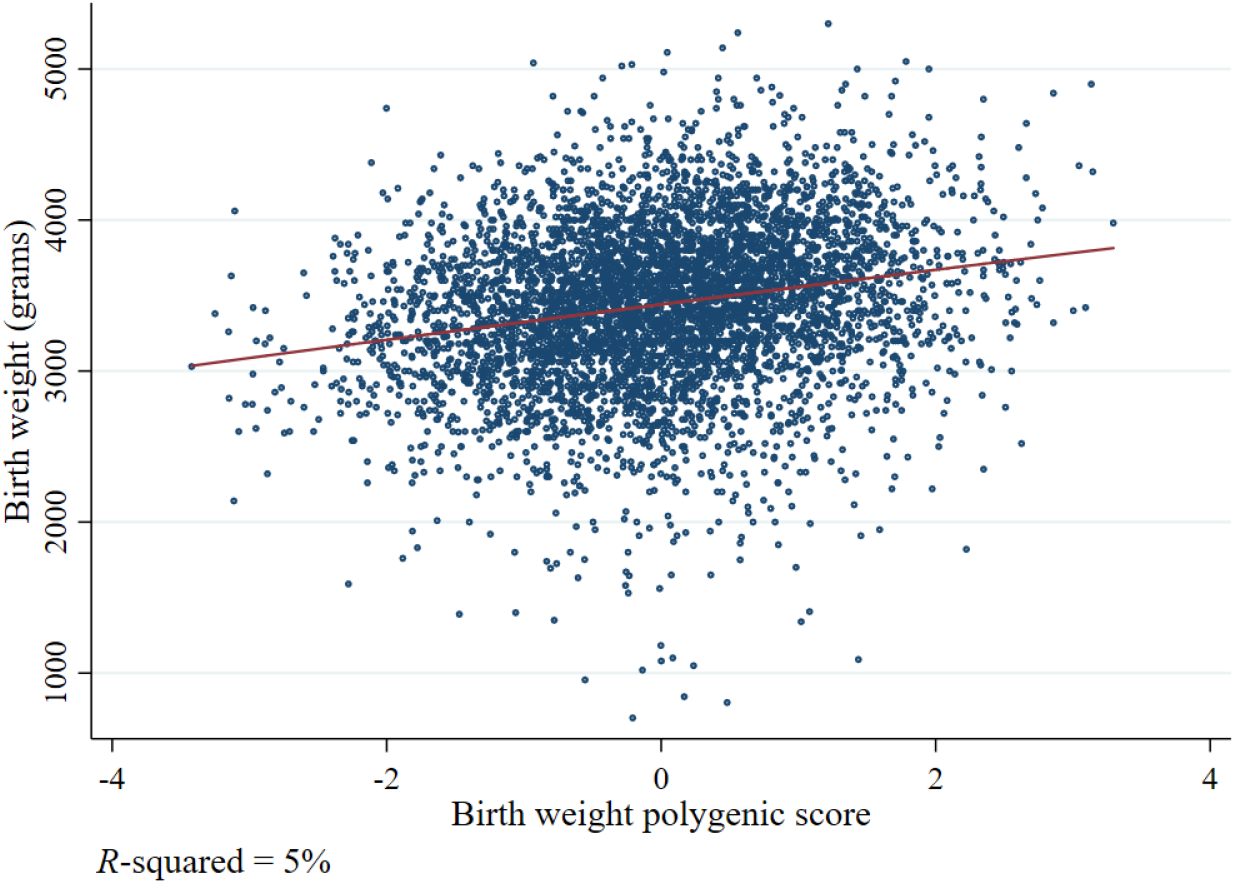
Results of a non-parametric regression of the actual birth weight of a child on the child’s birth weight polygenic score using the kernel density function for continuous covariates.

#### Instrumental variable

As we will elaborate upon in the next section, we instrument the number of cigarettes smoked per day with a SNP *of the mother* : the SNP rs1051730 which is located in the nicotine receptor gene CHRNA3. This genomic region located in the chromosome 15 cluster of virtually adjacent nicotinic receptor genes (CHRNA3, CHRNA5, and CHRNB4) was identified in all Genome-Wide Association Studies of smoking as a risk factor for the intensity of smoking defined by the number of cigarettes smoked per day (e.g., Furberg et al., 2010; Liu et al., 2019).

##### rs1051730

This variable is equal to 0 if the mother has no A(denine) nucleotide for SNP rs1051730, 1 if the mother has 1 A nucleotide and 2 if the mother has 2 A nucleotides. The adinine nucleotide of rs1051730 has consistently been associated with an increased number of cigarettes consumed per day. In our baseline sample, 45% of mothers do not carry any A nucleotides for this SNP, 44% have one A nucleotide, and the remaining 11% of the mother carry two risk alleles.

#### Control variables

Although we exploit random-like and exogenous variation that only influences the number of cigarettes smoked by the mother, we include some control variables for three reasons. First, we would like to compare our IV results to the results of an ordinary OLS regression with control variables. Second, the control variables act as a sensitivity analysis for our IV regression: since an instrumental variable should not be correlated with the control variables, including the control variables should not change the coefficient of interest. Third, including control variables that explain some of the variation in the outcome could help to increase precision. The control variables only involve characteristics of mother and grandmother, as these are the only variables possibly preceding offspring genetic endowments and birth weight.^8^ They include maternal birth weight, maternal age, maternal marital status, maternal education, maternal social class, and grandmother’s education (see Appendix A.4 for definitions and Table 1 for descriptive statistics). All control variables are included as (and when necessary transformed into) categorical variables with a dummy for each category, including a category for missing values (see Appendix A.4 for details). Finally, the first four principal components of the mother’s genetic relationship matrix are included to control for subtle forms of “population stratification”, i.e., systematic relationships between the prevalence of genetic variants and environments in subpopulations (Price et al., 2006; Rietveld et al., 2014).

## 3. Identification strategy

Our interest is in the effect of the number of cigarettes smoked per day during pregnancy (*C*_*M*_) on the child’s birth weight (*B*_*C*_). Note that we use the subscript *C* to denote the child’s variables and the subscript *M* to denote those of the mothers. A conventional OLS regression of the form

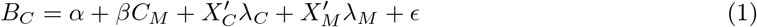

where *X*_*C*_ and *X*_*M*_ represent the child’s and maternal control variables as defined in section 2, respectively, is unlikely to produce an unbiased coefficient *β*. As already touched upon in the introduction, the number of cigarettes smoked per day *C*_*M*_ is subject to endogeneity concerns as a result of measurement error and omitted variables. To address the endogeneity of maternal smoking during pregnancy arising from omitted variables, we resort to an instrumental variable (IV) approach. In particular, we instrument the average number of cigarettes smoked per day *C*_*M*_ with the SNP rs1051730, which has been consistently linked with smoking intensity (e.g., Furberg et al., 2010; Liu et al., 2019) while being plausibly exogenous to offspring birth weight. The application of genetic instrumental variables is often referred to as Mendelian Randomization (Davey Smith, 2003; von Hinke et al., 2016).

In terms of equations, our identification strategy is therefore as follows. We use the following two equations:

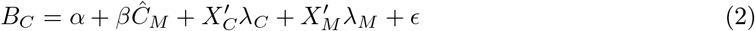

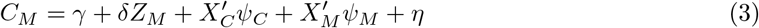

where *Z*_*M*_ denotes the maternal SNP rs1051730 used as instrumental variable and *ϵ* and *η* denote the error terms. A causal interpretation of the resulting two-stage least squares (2SLS, or Mendelian Randomization) estimates relies on three critical assumptions (see e.g., von Hinke et al., 2016), which we discuss in detail below.

First, our instrumental variable should have a strong effect on the endogenous variable of interest: *δ* ≠ 0. Several GWAS studies have robustly replicated the association between the SNP rs1051730 and smoking intensity (Furberg et al., 2010; Liu et al., 2010, 2019). The SNP is known colloquially among researchers as “Mr. Big” because of its consistently estimated large effect size.^9^ In our sample, as we will show below, the first stage estimate is positive and statistically significant, with the effective *F*-statistic values consistently in the range 15-25. Therefore, the relevance assumption seems to hold for our instrument (Olea and Pflueger, 2013; Andrews et al., 2019).

Second, our instrumental variable should be independent of confounding factors. An important advantage of using genetic variants as instruments is that they are randomly distributed at conception, conditional on population stratification variables, or, more stringent, parental genotype or family fixed effects. Therefore, the independence assumption is likely to hold when using SNPs as instruments (Davey Smith et al., 2007) in particular in our ALSPAC sample which is a very homogeneous sample from a relatively small geographic area. Moreover, we focus on mothers from European Ancestry only, and thus a correlation between smoking-related genetic variants and the error term of the outcome equation (2) is unlikely. Although not a proof, it is reassuring that our instrumental variable is not correlated to any of the background variables of the mother or child (see section 4). Moreover, importantly, the SNP is also not correlated to the child’s birth weight polygenic score, suggesting that our estimates are not biased by a possible gene-environment correlation (*rGE*).^10^

Third, our instrumental variable is only allowed to impact the child’s birth weight through smoking. The so-called exclusion restriction is the most challenging one when employing genetic variants as instrumental variables as typically the biological function of a certain gene is not completely known and one cannot rule out so-called “biological pleiotropy”: i.e. the same SNP affecting multiple outcomes (Lawlor et al., 2017). The main advantage of using SNP rs1051730 is that its biological function is well understood. In particular, it is known to cause an amino acid change in the alpha-5 subunit of the nicotinic receptors, and experiments have found that this change alters the responsiveness of the nicotinic receptors to nicotine (Bierut and Cesarini, 2015). Hence, the SNP relates to nicotine dependence and the channel through which the SNP affects children’s outcomes is plausibly maternal smoking. In accordance with this mechanism, when we stratify the sample according to whether the mother smoked during pregnancy (e.g., Van Kippersluis and Rietveld, 2018a,b), we do find a strong association between our IV and birth weight among smoking mothers, whereas there is no significant relationship among non-smoking mothers. Even though the stratification into these subgroups may be endogenous,^11^ we believe this result (shown in section 4) is reassuring and suggests that the exclusion restriction holds in our model.

Although we believe that instrumenting the average number of cigarettes smoked per day during pregnancy with our genetic instrument goes a long way in tackling the endogeneity of maternal smoking, the possible misreporting of smoking still affects our first stage and hence 2SLS results.^12^Therefore, our main analyses are based upon the reduced form of the 2SLS approach (i.e., the intention-to-treat effect of the SNP on offspring birth weight):

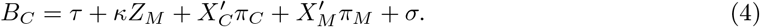

The reduced form does not rely on the possibly biased self-reported smoking measures, and a significant coefficient *κ* is a necessary condition for a causal effect of the instrumented variable (maternal smoking) on the outcome (birth weight). Still, we also present 2SLS results using various measures of maternal smoking in the first stage, in order to assess what a reasonable first stage (denominator) would be to scale our reduced form (numerator) effects into a plausible 2SLS estimate.

In order to assess the interaction between genes – as measured by the child’s PGS for birth weight – and maternal smoking, we amend our regressions (2), (3) and (4) by including a linear interaction term between the maternal SNP and the child’s polygenic score.^13^Additionally, we estimate models (2), (3) and (4) separately for four quartiles of the PGS for birth weight to allow for possible non-linearities in the interaction between the child’s genes and maternal smoking.

## 4. Results

In this section we present our main empirical results. We start by discussing the results of the conventional OLS regressions. Thereafter, we discuss the IV results using the maternal SNP as instrumental variable and assuming homogeneous effects. Finally, we turn our attention to the heterogeneity analyses using interaction terms between the PGS for birth weight and maternal smoking and using stratification by quartiles of the child’s PGS for birth weight.

### 4.1. OLS regression results

Table 2 present the results of OLS regressions in which the child’s birth weight is the outcome variable, and the number of cigarettes smoked per day is the explanatory variable. All specifications consistently show a statistically significant association between the number of daily cigarettes smoked by the mother and the child’s birth weight, with every daily cigarette smoked associated with a reduction between 8 and 15 grams in birth weight. The coefficient of the PGS for birth weight is stable across specifications, and statistically significant at the 1% level. Every standard deviation increase in the PGS increases one’s birth weight by around 108-120 grams, consistent with the findings of e.g. Trejo (2020). Similar patterns are found when using the binary indicator for whether the mother smoked or not during pregnancy. These results can be found in Appendix A.5.

**Table 2.** Results of the OLS regressions explaining the child’s birth weight. Coefficients are displayed with robust standard errors in parentheses. Column 1-3 use the self-reported smoking measure and Column 4-6 the self-reported smoking measure corrected using cotinine levels. All regressions correct for genetic relatedness among the mothers using the first four principal components of the genetic relationship matrix.

|  | (1) | (2) | (3) | (4) | (5) | (6) |
| --- | --- | --- | --- | --- | --- | --- |
| #Cigarettes smoked per day | -14.8**<br>(1.3) | -12.7**<br>(1.3) | -13.4**<br>(1.7) |  |  |  |
| #Cigarettes smoked per day (adjusted) |  |  |  | -12.0**<br>(1.3) | -8.3**<br>(1.5) | -9.4**<br>(1.8) |
| PGS of birth weight |  |  | 108.5**<br>(6.9) |  |  | 118.7**<br>(10.0) |
| Control variables | No | Yes | Yes | No | Yes | Yes |
| *R*-squared | 0.019 | 0.059 | 0.101 | 0.021 | 0.069 | 0.128 |
| *N* | 7,598 | 7,598 | 5,006 | 3,784 | 3,784 | 2,408 |

+ $p < 0.1$ , \* $p < 0.05$ , \*\* $p < 0.01$

An interesting result stemming from Table 2 is that the correction of the self-reported smoking measure using cotinine levels lowers the coefficients from −13.4 to −9.4 (and from −144.8 to −57.6 on the extensive margin, see Appendix A.5).^14^ Analyses presented in Appendix A.6 may explain why measurement error in the smoking measure biases the effect away from zero. These analyses show that mothers who smoke according to their cotinine level but who do not report to be smokers, are more similar to the non-smoking mothers than to self-reported smoking mothers. One possible explanation is that the mothers who smoke and choose not to report are more aware of the dangers of smoking and therefore more reluctant to report smoking. However, while being better informed about smoking they might be more careful overall during their pregnancy compared with mothers who report to be smoking.^15^

### 4.2. IV regression results assuming homogeneity

#### First stage

We start by analyzing the results of the first stage of our Instrumental Variables strategy in Table 3. The most important result is that our instrument has a significant and strong impact on maternal smoking intensity during pregnancy. A risk allele is estimated to increase the number of cigarettes per day by around half a cigarette. Hence, women with two risky alleles (about 11% of the sample) smoke on average 1 extra cigarette per day, an increase of 18% relative to the mean of our adjusted measure of the number of cigarettes smoked per day. In each specification, the coefficient for the SNP rs1051730 is significant at the 1% significance level, and the effective *F*-statistic is at least 15 in each specification. The effective *F*-statistic (Olea and Pflueger, 2013) drops the assumption of homoskedasticity and it is therefore considered more appropriate than the standard *F*-statistic. One can also note that the coefficient is larger (albeit with larger standard errors) in the specification in which we only include mothers with a measure of cotinine (Columns 3 and 4). These differences suggest that our adjustment of the number of cigarettes indeed reduces measurement error in our treatment variable.

**Table 3.** Results of the OLS (first stage) regressions explaining smoking. Coefficients are displayed with robust standard errors in parentheses. Column 1-2 use the self-reported smoking measure, and Column 3-4 the self-reported smoking measure corrected using cotinine levels. All regressions correct for genetic relatedness among the mothers using the first four principal components of the genetic relationship matrix.

|  | #Cigarettes smoked<br>per day |  | #Cigarettes smoked<br>per day (adjusted) |  |
| --- | --- | --- | --- | --- |
|  | (1) | (2) | (3) | (4) |
| rs1051730 | 0.5**<br>(0.1) | 0.4**<br>(0.1) | 0.6**<br>(0.2) | 0.6**<br>(0.1) |
| Control variables | No | Yes | No | Yes |
| *R*-squared | 0.004 | 0.132 | 0.005 | 0.005 |
| Effective *F*-statistic | 25.3 | 15.6 | 21.4 | 27.2 |
| *N* | 7,598 | 7,598 | 3,784 | 3,784 |

<sup>+</sup> $p < 0.1$ , \* $p < 0.05$ , \*\* $p < 0.01$

#### Reduced form

Column 1-2 in Table 4 show the reduced forms in our baseline sample 1. The coefficients imply that having one A allele of the SNP rs1051730 decreases offspring birth weight by on average 17 grams. The effect is statistically significant at the 10% level. The reduced form in the sample of 8,399 mothers – where we do not impose the baseline restriction of observing maternal smoking – reveals a reduced form of −19g, significant at the 5% level (not shown). We therefore conclude that our reduced form result supports the interpretation that maternal smoking during pregnancy causally reduces offspring birth weight. Reassuringly, the result remains stable when we add the maternal control variables. These results further support our assumption that this particular SNP is likely to be as-good-as randomly assigned in the relatively homogeneous population residing in the Avon area from which the ALSPAC participants come from.

**Table 4.** Results of the OLS (reduced form) regressions explaining birth weight. Coefficients are displayed with robust standard errors in parentheses. All regressions correct for genetic relatedness among the mothers using the first four principal components of the genetic relationship matrix.

|  | Baseline Sample 1 |  | Smokers<br>(Cotinine > 100 ng/ml) |  | Non-smokers<br>(Cotinine < 100 ng/ml) |  |
| --- | --- | --- | --- | --- | --- | --- |
|  | (1) | (2) | (3) | (4) | (5) | (6) |
| rs1051730 | -16.7+<br>(9.6) | -16.7+<br>(9.5) | -42.9<br>(29.3) | -66.9*<br>(29.4) | 8.5<br>(17.8) | 9.3<br>(17.6) |
| Control variables | No | Yes | No | Yes | No | Yes |
| *R*-squared | 0.001 | 0.040 | 0.005 | 0.124 | 0.002 | 0.074 |
| *N* | 7,598 | 7,598 | 828 | 828 | 2,014 | 2,014 |

<sup>+</sup> $p < 0.1$ , \* $p < 0.05$ , \*\* $p < 0.01$

In order to investigate the independence assumption more formally, we assess the relationship of the maternal SNP with background characteristics in Table A7 in Appendix A.7. We conclude that the instrument is uncorrelated with the analyzed characteristics of the mother and grandmother.

In order to gauge the validity of the exclusion restriction of our instrumental variable, we evaluated if the SNP has any effect on the outcomes of mothers who should <u>not</u> be affected by the SNP. Non-smokers constitute such a sample, as the SNP rs1051730 is associated with the intensity of smoking. If the SNP has an impact on birth weight among children of mothers who did not smoke during pregnancy, then it suggests that there must be at least one other pathway other than smoking through which the instrument affects offspring birth weight (Van Kippersluis and Rietveld, 2018a,b). The existence of such a pathway would violate the exclusion restriction.

In Columns 3-6 of Table 4, we estimate the reduced form in stratified subsamples based on the cotinine threshold we previously defined for active smoking.^16^ Columns 3-4 indicate that the relationship between the SNP rs1051730 and birth weight is negative (and statistically significant in Column 4) among smoking mothers, whereas Columns 5-6 indicate that this relationship is positive, close to zero and non-significant among non-smoking mothers. This result supports the idea that the SNP impacts birth weight solely through smoking intensity and not through other pathways.

We acknowledge that these placebo tests are not sufficient to validate our exclusion restriction for at least two reasons. First, rejecting a non-zero effect among a group that should not be affected by the IV is obviously not evidence for a precise zero direct effect among the subgroup that is affected. Hence this test can never validate the exclusion restriction. Second, the stratification into smokers and non-smokers itself could naturally be endogenous to the IV. This implies that our group of mothers who do not smoke might be a slightly selected group of women who despite having this SNP decided not to smoke during pregnancy. It could be that this endogenous stratification biases the effect of the SNP on child’s outcomes downwards. In Appendix A.8, we show however that the distribution of the effect allele between smokers and non-smokers is not significantly different. Therefore, we conclude that selection into the group of smokers or non-smokers based on the SNP is not likely to be a major factor of concern. In sum, the absence of any effect among the group of non-smoking mothers in Column 3-6 of Table 4 is consistent with the validity of our exclusion restriction, and the striking difference in sign and magnitude of the effect of the SNP on the child’s outcome between smokers and non-smokers supports a causal interpretation of the intensity of maternal smoking during pregnancy on birth weight of the offspring.

#### 2SLS

Having provided evidence in support of the IV assumptions, and in order to interpret the magnitude of the reduced form effects in terms of number of cigarettes, we now move to the 2SLS results in Table 5. There are several patterns that emerge. First, all the IV estimates point estimates are remarkably stable, regardless of the measure of smoking one utilizes. The estimates imply that one extra cigarette per day reduces offspring birth weight by around 36-40 grams.

**Table 5.** Results of the 2SLS regression explaining birth weight. Coefficients are displayed with robust standard errors in parentheses. All regressions correct for genetic relatedness among the mothers using the first four principal components of the genetic relationship matrix. Columns and *F*-statistic as in Table 4.

|  | (1) | (2) | (3) | (4) |
| --- | --- | --- | --- | --- |
| #Cigarettes smoked per day | -36.7+<br>(21.1) | -37.9+<br>(21.7) |  |  |
| #Cigarettes smoked per day (adjusted) |  |  | -36.5+<br>(21.4) | -38.5<br>(23.9) |
| Control variables | No | Yes | No | Yes |
| Effective $F$-statistic | 25.3 | 15.6 | 21.4 | 27.2 |
| $N$ | 7,598 | 7,598 | 3,784 | 3,784 |

+ $p < 0.1$ , \* $p < 0.05$ , \*\* $p < 0.01$

Second, consistent with the validity of the independence assumption, the point estimates are hardly affected by the inclusion of control variables. If anything, in contrast with the OLS estimates from section 4.1, correction of the number of cigarettes smoked per day based on cotinine levels slightly increases rather than decreases the magnitude of the estimates in absolute value. The differences between the unadjusted and adjusted measures are however not statistically significant.

Third, and strikingly, the coefficients are considerably larger than in the baseline OLS regressions, although the confidence intervals span values from close to 0 to almost 75 grams and thus cover the OLS estimates. The fact that the IV point estimates are larger than the OLS estimates is somewhat surprising if one considers the reduction in the coefficient by adding control variables in the OLS regression. This would suggest that omitted variables bias would bias the OLS estimates upwards, not downwards. However, the larger IV point estimates could possibly be explained by the IV results being less prone to random measurement error. An additional explanation could be that the Local Average Treatment Effect (LATE) among compliers (i.e., the mothers on the margin induced to smoke by their genotype) is larger in magnitude than the Average Treatment Effect on the Treated (ATT) that OLS seeks to estimate.

Finally, it could be that there still exists measurement error in our adjusted measures for the number of cigarettes smoked per day. This would attenuate the OLS estimates, yet plausibly leads to an overestimation of the IV estimates.^17^ In section A.9 in the Appendix, we drop the assumption that people who reported a positive number of cigarettes reported correctly. Rather, we assume that they under-reported this number. However, even when on average mothers under-report the number of cigarettes smoked per day by 10, the IV point estimate is still somewhat larger than the OLS estimate and it is estimated to be around −20 grams for every daily cigarette smoked during pregnancy.

Multiplying these IV estimates by the average number of cigarettes smoked per day among smoking mothers (around 10), would imply that offspring birth weight among smoking mothers is around 200-400 grams lower than among non-smoking mothers. While these estimates are surrounded by pretty large uncertainty, and even though the values are not not outrageously higher than existing estimates in the literature (Zheng et al., 2016; Kataoka et al., 2018), they do seem large compared to other well-known risk factors for birth weight such as nutrition (e.g., Almond et al., 2011; Barber and Gertler, 2008), and stress (Aizer, 2011; Currie and Rossin-Slater, 2013; Black et al., 2016; Persson and Rossin-Slater, 2018; Doyle et al., 2020). Therefore, since the IV estimates rely on an imperfect measure of smoking in the first stage, we report our heterogeneity results on basis of the reduced form only in the next subsection.

### 4.3. Reduced form regression results assuming heterogeneity

Our final analysis in the ALSPAC sample seeks to assess whether the effect of maternal smoking during pregnancy affects children differently with respect to their genetic propensity to be born with low or high birth weight. Table 6 presents the reduced form regression in which we interact the maternal SNP rs1051730 with the child’s birth weight polygenic score. Note that we multiply the child’s PGS by −1, to make sure both variables have negative effects on the outcome variable, facilitating the interpretation of the interaction term.

**Table 6.** Results of the OLS (reduced form) regression explaining birth weight. Coefficients are displayed with robust standard errors in parentheses. All regressions correct for genetic relatedness among the mothers using the first four principal components of the genetic relationship matrix.

|  | (1) | (2) |
| --- | --- | --- |
| rs1051730 | -1.0<br>(10.9) | -3.0<br>(10.8) |
| (-1) Birth weight PGS (child) | -114.2**<br>(10.1) | -103.6**<br>(10.0) |
| rs1051730 × (-1) Birth weight PGS (child) | -2.8<br>(10.5) | -5.3<br>(10.4) |
| Control variables | No | Yes |
| *R*-squared | 0.047 | 0.077 |
| *N* | 5,006 | 5,006 |

+ $p < 0.1$ , \* $p < 0.05$ , \*\* $p < 0.01$

In this model, both the direct effect of the SNP as well as the interaction term are non-significant, with both coefficients relatively close to zero. Apparently, the drop in sample size because of the restriction to include mother-child pairs for which the child’s genotypes are observed causes the main effect of the SNP to be insignificant and smaller in magnitude compared to the estimate in Table 4. The effect of the PGS for birth weight is again significant at the 1% level with a similar coefficient compared with the OLS results. Even though the magnitude of the interaction term is very small (especially compared to the main effect of the PGS), concluding the absence of meaningful interactions between genes and maternal smoking in causing birth weight would be premature for two reasons.

First, Table 6 only presents the rather restrictive form of a multiplicative linear interaction, whereas non-linear interaction could also be present. In order to investigate possible non-linear interactions, we run the regression from Table 4 separately for subsamples based on quartiles of the distribution of the child’s PGS of birth weight. The full set of regression results is available in Appendix A.10, but a graphical summary of the results can be found in Figure 2 (left-panel). The Figure again shows no evidence of meaningful heterogeneity in the effect of maternal smoking on the child’s birth weight.

**Fig 2.**
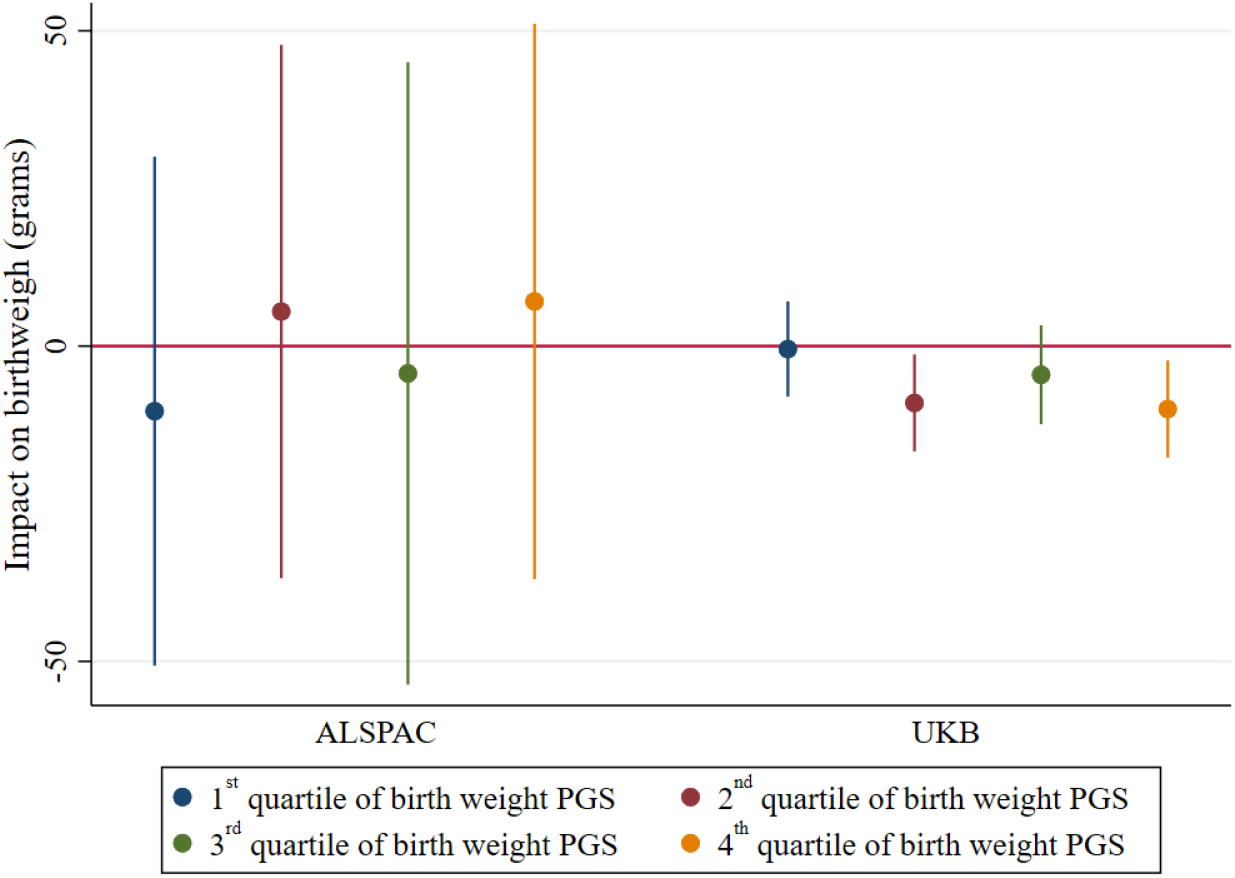
Results of the OLS (reduced form) regressions explaining birth weight for subsamples in ALSPAC and UK Biobank based on quartiles of the children’s PGS of birth weight. The coefficient of each regression is plotted together with its robust 95% confidence interval. In the regressions, the full set of control variables is included. More details about these regressions are available in Appendix A.10.

A second reason why we cannot firmly state the absence of *G* × *E* interactions is that we may simply lack statistical power to detect it. The confidence intervals of the reduced form effect among the four quartiles of the PGS in Figure 2 (left panel) are rather wide, and the first stage results in these samples are below conventional thresholds (see Table A11 in Appendix A.10). Therefore, even though these results provide tentative evidence that strong interactions between a child’s PGS and maternal smoking do not exist, they are not conclusive. For this reason, we replicate these reduced form results in section 5 using the much larger UK Biobank to assess whether the absence of evidence can be attributed to low statistical power.

## 5. Replication

In this section, we provide the results of the replication analysis using data from the UK Biobank (UKB).

### 5.1. Data and variables

The UKB is a UK data set collected with the aim of improving the prevention, diagnosis and treatment of a wide range of serious and life-threatening illnesses. The UKB recruited approximately 500,000 people aged between 40-69 years in 2006-2010 from across the UK to take part in this project. They have undergone measurements, and provided detailed information about themselves as well as blood, urine and saliva samples. Genotyping has been undertaken on all 500,000 participants. More information on the genetic section of the UKB can be found in (Bycroft et al., 2017).

The UKB does not have the level of measurement detail as ALSPAC has. For our replication purposes, the main data limitations are as follows. First, the UKB does not provide the average number of cigarettes smoked during pregnancy nor a cotinine measure for cross validation. Second, it only has genetic information on the individual itself, and no family members were targetly genotyped.^18^ Finally, no information about the socio-economic background of the mothers is available, ruling out the possibility to add control variables to the model. Nevertheless, the sample size of the UKB is much larger than that of ALSPAC enabling us to assess whether lack of statistical power is driving the absence of evidence of significant *G* × *E* interactions in birth weight in the main analyses with the following variables that are available.

#### Birth weight

Participants were asked to enter their own birth weight. Given that all individuals were alive past 1 year old, no outliers were removed. The mean birth weight is 3,325g and the standard deviation is 665g.

#### Maternal smoking around birth

Participants were asked the following question “Did your mother smoke regularly around the time when you were born?”. They could reply with yes or no. This variable was collected from all participants except from those who indicated they were adopted as a child. 30% of respondents answered “yes”.

#### rs1051730

This variable is equal to 0 if the individual has no A’s nucleotide for SNP rs1051730, 1 if he or she has 1 A nucleotide and 2 if he or she has 2 A nucleotides. In the sample, around 45% do not have any A nucleotides, 44% has one A nucleotide, and the remaining 11% has two A alleles. Note that this distribution in exactly the same as in ALSPAC. More details on the quality control of the genetic data can be found in Appendix A.2. Importantly, in the UKB analyses, we rely on the *individual’s* SNP rather than the preferred *maternal* SNP to instrument the *maternal* smoking decision during pregnancy. Essentially, this approach uses the child’s SNP as a proxy for the maternal SNP, which in turn is an instrumental variable for maternal smoking during pregnancy. While this may seem far-fetched, Yang et al. (2019) provide compelling evidence that with this strategy it is possible to detect causal effects of maternal smoking on offspring birth weight. It relies on (i) Mendel’s law that children randomly inherit 50% of the maternal genes, and (ii) the same exclusion restriction as before that the single causal channel of the nicotine receptor gene is the intensity of smoking. Since unborn children clearly did not make any smoking decisions, it is plausible that the only reason why the child’s SNP affects his/her birth weight is through maternal smoking. While obviously using a child’s SNP as a proxy for his/her mother’s SNP is introducing some measurement error, the additional power gained by the sample size enables us to replicate our main findings in ALSPAC using a larger sample.

#### Birth weight polygenic score

We use the summary statistics of Warrington et al. (2019) depleted of the UKB to retrieve the relevant SNPs with their weights. We construct the PGS using the software LDPred (Vilhjalmsson et al., 2015). The predictive power of the PGS is 1.6%, and individuals in the lowest decile of the PGS distribution have on average a 300 grams lower birth weight than those in the highest decile. The *R*-squared of the PGS is lower in the UKB than it is in ALSPAC since the UKB was the largest sample in the GWAS. However, since the UKB is our hold-out cohort for this analysis, we are forced to deplete the GWAS summary statistics from the UKB to avoid overfitting. More details about the construction of the PGS can be found in Appendix A.3.

#### Principal components

We add the first 20 principal components of the genetic relationship matrix, as provided by the UKB, to control for subtle population stratification (Price et al., 2006; Rietveld et al., 2014). The inclusion of these control variables to our models does not qualitatively influence any of our results.

### 5.2. Results

In this subsection we present our IV results with and without assessing treatment effect heterogeneity. For reasons of brevity, the OLS results are presented in Appendix A.5. Since the intensity of smoking during pregnancy was not reported in the UKB, it is not possible to replicate our first stage results from ALSPAC directly. We can only check the relationship between the instrument and the smoking status of the mother on the extensive margin (smoking vs. not smoking). However, given the nature of the biological mechanism triggered by the SNP rs1071730, this effect is expected to be much weaker. Nonetheless, if we run this first stage regression (not shown), the coefficient is 0.004 and statistically significant at the 5% significance level. This results suggests that for each effect allele an individual is carrying the probability of the mother smoking around birth is 0.4% higher.

In Table 7 we present the reduced form effect of our IV on the birth weight of the respondent, for the full sample and stratified subsamples by smoking status of the mother. Some respondents did not report the smoking status of their mother, and therefore the subsamples do not sum up to the full sample. In the full sample, one effect allele of the SNP decreases birth weight by around 6 grams on average (Column 1). However, the effect is −17 grams in the subsample of individuals with a smoking mother (Column 2) and non-significant in the subsample of individuals with a non-smoking mother (Column 3).^19^

**Table 7.** Results of the OLS (reduced forms) regressions explaining birth weight. Coefficients are displayed with robust standard errors in parentheses. All regressions correct for genetic relatedness using the first twenty principal components of the genetic relationship matrix.

|  | Full sample | Smoking Mothers | Non-smoking Mothers | Full sample |
| --- | --- | --- | --- | --- |
|  | (1) | (2) | (3) | (4) |
| rs1051730 | -5.7** | -17.3** | -1.3 | -6.1** |
|  | (2.0) | (4.0) | (2.4) | (2.0) |
| (-1) Birth weight PGS (child) |  |  |  | -83.0** |
|  |  |  |  | (1.8) |
| rs1051730 × (-1) Birth weight PGS (child) | | | | 1.0 |
|  |  |  |  | (2.0) |
| *R*-squared | 0.001 | 0.001 | 0.001 | 0.016 |
| *N* | 256,702 | 67,915 | 160,530 | 256,702 |

+ $p < 0.1$ , \* $p < 0.05$ , \*\* $p < 0.01$

We cannot readily compare the reduced forms in the UKB and ALSPAC because we use the child’s SNP (as a proxy-instrument) in UKB and the maternal SNP in ALSPAC. Still, given the random inheritance of SNPs we can gauge the similarity of effect sizes. In ALSPAC, the correlation between rs1051730 among mothers and their children is 50.7%, and an additional A allele of the mother for this SNP is associated with a 16.7 grams decrease of the offspring birth weight (see Table 4). In the UKB, an additional A allele of the child for this SNP is associated with a 5.7 grams decrease of birth weight. Hence, we may expect a partial correlation between the A allele of the child and birth weight in the UKB of −16.7 × 0.507 = −8.5 grams. It is reassuring that this number is within the confidence interval of what we find in the UKB.

Given that we do not have a measure of the average number of cigarettes smoked during pregnancy in the UKB data set, we rely on the first stage estimated in ALSPAC (0.6, see Columns 3-4 of Table 3) in order to infer an IV estimate. A manually computed Two-Sample Two Stage Least Squares (TS2SLS) estimate would be −5.7 × 2*/*0.6 = −19 grams. Therefore, a rough estimate is that an average daily cigarette reduces birth weight by around 20 grams. This effect is somewhat smaller compared with what we find in our main analysis in ALSPAC, but pretty close to the estimates we find in ALSPAC after taking into account measurement error in self-reported smoking.

The results in the previous subsection indicate that the UKB is a suitable (second-best) sample to estimate the causal effect of maternal smoking on offspring birth weight. However, our main interest is in assessing the presence of heterogeneity of the treatment effect in the UKB. Column 4 of Table 7 presents the reduced form results including the interaction. The main effects of the SNP and the PGS are significant, but as in ALSPAC the interaction term is close to zero. However, in contrast with the ALSPAC results, the standard error of the interaction term is now also very small allowing us to rule out interaction terms outside the (−1 to 3) grams bandwidth with 95% confidence. Further, the stratification of the reduced form effect by quartiles of the PGS shown in Figure 2 (right panel) further supports the claim of no meaningful interaction effect between the birth weight PGS and maternal smoking. The stratification of the reduced form effect by quartiles of the PGS in Figure 2 shows remarkably narrower confidence intervals than in ALSPAC but similarly shows no clearly discernible patterns. Moreover, the effect estimates across quartiles are not significantly different from each other. Overall, we take this as evidence that statistical power was not the main driver of the lack of evidence for an interaction effect in ALSPAC.

## 6. Discussion and conclusion

In this study we show that maternal smoking during pregnancy as well as one’s genetic predisposition contribute to offspring birth weight. In particular, we find that a 1-cigarette increase in the average number of cigarettes smoked per day during pregnacy reduces birth weight, with point estimates varying from around 20 grams (UKB and ALSPAC with underreporting correction) to 40 grams (ALSPAC without underreporting correction). A one standard deviation increase in the polygenic score increases birth weight by around 80 (UKB) to 120 (ALSPAC) grams. Despite the strong and well-established main effects, our results suggest that there is no meaningful interaction effect among these two drivers of birth weight. Hence, maternal smoking does not exacerbate genetic inequalities, and even a high genetic predisposition to birth weight cannot cushion against the damaging environmental exposure of maternal smoking during pregnancy.

These findings contribute to the literature on the genetic and environmental determinants of offspring birth weight, an important barometer of pregnancy outcomes (Trejo, 2020). By using innovative identification strategies that simultaneously address measurement error and endogeneity concerns, our results re-emphasize the damaging effects of smoking during pregnancy. Moreover, we highlight the role of genetic endowments implied in birth weight. Our main contribution is to show the absence of heterogeneity in the effect of maternal smoking according to the child’s genotype. Hence, our results suggest that for birth weight both nature (here measured by the PGS for birth weight) as well as nurture (here measured by maternal smoking during pregnancy) impact birth weight, but we do not find evidence of meaningful interactions between the two.

Three main limitations should be acknowledged. First, birth weight is not the only relevant early-childhood outcome, and many genetic as well as environmental drivers could influence later life outcomes without affecting birth weight (Conti et al., 2018). Therefore, we cannot rule out meaningful interactions between genes and maternal smoking decisions for other life outcomes. In auxiliary analyses we investigate the effect on weeks of gestation and whether the child is alive 1 year after birth in ALSPAC, but we do not detect meaningful main effects or interactions (results available upon request from the authors). Future studies may want to analyze whether such effects exist as well as their size in sufficiently powered analyses.

Second, it has been argued that maternal smoking before pregnancy may affect oocyte quality and thereby birth outcomes (e.g., Oyesanya et al., 1995). If true, this would represent an effect of maternal smoking *before* pregnancy and our estimates should be interpreted as the effect of all pre-natal smoking exposure rather than just exposure to smoking during pregnancy.

Finally, the PGS for birth weight is not a perfect measure of the child’s genetic endowments for birth weight. It only captures common SNPs and not rare variants or other genetic variation across humans. Moreover, the PGS is constructed on basis of a Genome-Wide Association Study (GWAS) that is pooling various cohorts exposed to different environmental conditions. Mills et al. (2020) therefore argue that a PGS may actually over-weigh SNPs that are particularly resilient to environmental conditions, which would go against finding a significant *G* × *E* effect when using a PGS. The limited sample size and sparsity on control variables of the GWAS discovery analysis may also introduce measurement error in PGSs, and possibly a bias as a result of the omission of parental genotype from the GWAS. If parental genes influence the child’s birth weight through other mechanisms than purely inheritance of genes, these other mechanisms would be captured in the PGS too. Nevertheless, within-family predictions with the PGS however shows that bias is not a major concern for the birth weight PGS, and interactions with other (endogenous) maternal characteristics such as BMI have been found (Trejo, 2020).

What could however be an issue is that the predictive power of the PGS for birth weight does not fully explain the heritability of birth weight (Warrington et al., 2019). With a more predictive PGS, small (interaction) effects may become statistically detectable, and hence it would be interesting to repeat our estimations as soon as results of a new GWAS on birth weight become available. We caution that these results are not to be expected in the next few years, as we draw on a very recent GWAS on birth weight to construct our polygenic scores and it usually takes several years for the GWAS sample size has been sufficiently expanded to conduct a new GWAS. Moreover, related to the previous point, the inclusion of additional genotyped samples in the GWAS meta-analysis may increase the chances that SNPs are over-weighted in the PGS that are particularly resilient to environmental conditions.

Therefore, we believe that the present study contains the most comprehensive analysis of the impact of the heterogeneous impact of maternal smoking on birth weight by genetic endowments which is achievable today. As such, it can also serve as a template for future *G* × *E* studies on related exposures and outcomes.

## Data Availability

The data can be obtained by filing a request directly to the University of Bristol (https://proposals.epi.bristol.ac.uk/)

## A. Appendix

### A.1. Cotinine

Despite cotinine being commonly used as a biomarker to validate a self-reported smoking status, the choice of the cotinine cut-off value for distinguishing true smokers from non-smokers differs among studies (Kim, 2016). There are three main reasons for this heterogeneity. First, cotinine levels vary according to how much nicotine one inhales, how much nicotine is in one cigarette and the pattern of use. In addition, the processing rates of cotinine vary between individuals, in particular, different processing rates were found for different ethnic groups (Benowitz et al., 2009). Finally, there has been a drop in advocated cut-offs over the last 20 years (Kim, 2016) which can be linked to an increase in regulations and awareness of the general public for the dangers of nicotine. Given this heterogeneity, one would expect that the distribution of cotinine is bimodal, with different peaks for smokers and non-smokers and variation around the peaks. For smokers, the distribution reflects primarily active smoking whereas for non-smokers the distribution mostly reflects passive smoking. When selecting a threshold, there is naturally a trade-off between type I and type II errors.

Kim (2016) reviews the literature and claims that urine cotinine cut-off values ranging from 50-200 ng/ml are commonly used to validate self-reported smoking status. Still, some of the reviewed studies rely on self-reported measures of smoking which might be biased and therefore bias the optimal thresholds selected. In order to select our threshold, we mainly focus on two studies which, in our view, manage to overcome this issue. Hoffmann et al. (1984) expose individuals to various degrees of smoke while measuring their urine, saliva and serum cotinine. In the most intense treatment, where people were exposed to the smoke of 4 cigarettes for 300 minutes, cotinine levels in urine were as high as 55 ng/ml. de Weerd et al. (2002) rely on a sample exclusively composed of people who are self-reported passive and active smokers. This reduced the scope of misreporting, given that people are likely to under-report but not likely to over-report smoking. On average, passive smokers had urine cotinine levels ranging from 34 to 73 ng/ml, while active smokers who smoked 1-9 cigarettes a day had between 411 to 713 ng/ml. The minimum cotinine level for active smokers was 71 ng/ml.

It is clear from the literature that an uncontroversial threshold simply does not exist. Nonetheless, we still believe that using this measure has many advantages as long as we use a threshold that minimizes type I errors. Plausibly, if women misreport their cigarette consumption during pregnancy, they could under-report, but are very unlikely to over-report. Therefore, we choose to re-classify women with really high cotinine levels during pregnancy as active smokers, where we use a reasonable yet conservative threshold of 100 ng/ml. Table A1 presents the mean cotinine levels by self-reported smoking status. Note that 40% of the self-reported non-smokers had 0 ng/ml in their urine and 85% had less than 100ng/ml. Only 4% of the self-reported smokers has 0 ng/ml and 12% had less than 100ng/ml (not shown) of cotinine in their urine. As expected, the distribution for the self-reported non smokers peaks at 0ng/ml, with a lot of probability mass for small amounts of cotinine. As such, we only re-classified real outliers as smokers.

**Table A1.**
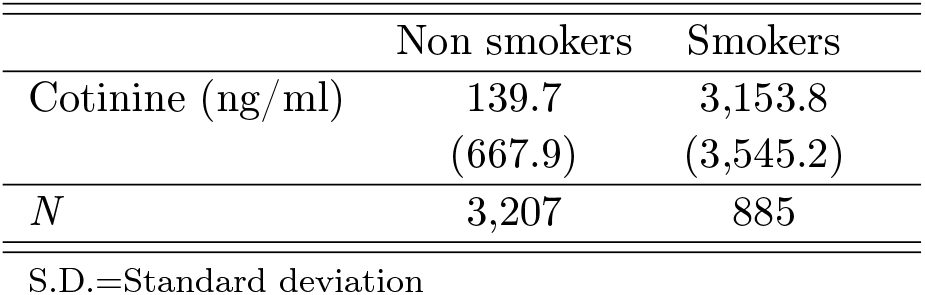
Cotinine levels (means with standard deviations in parentheses) in the first trimester by self-reported smoking status

### A.2. Quality control genetic data

#### ALSPAC children

ALSPAC children were genotyped using the Illumina HumanHap550 quad chip genotyping platforms by 23andMe subcontracting the Wellcome Trust Sanger Institute, Cambridge, UK and the Laboratory Corporation of America, Burlington, NC, US. The resulting raw genome-wide data were subjected to standard quality control methods. Individuals were excluded on the basis of gender mismatches; minimal or excessive heterozygosity; disproportionate levels of individual miss-ingness (>3%) and insufficient sample replication (IBD < 0.8). Population stratification was assessed by multidimensional scaling analysis and compared with Hapmap II (release 22) European descent (CEU), Han Chinese, Japanese and Yoruba reference populations. All individuals with non-European ancestry were removed. SNPs with a minor allele frequency of < 1%, a call rate of < 95% or evidence for violations of Hardy-Weinberg equilibrium (*p* < 5 ×10^−7^) were removed. Cryptic relatedness was measured as proportion of Identity by Descent (IBD > 0.1). Related subjects that passed all other quality control thresholds were retained during subsequent phasing and imputation. 9,115 subjects and 500,527 SNPs passed these quality control filters. The imputation was performed using the IMPUTE v2.2.2 software using the 1000 genomes phase 1 version 3 reference panel. Monomorphic or singleton SNPs were excluded.

#### ALSPAC mothers

ALSPAC mothers were genotyped using the Illumina human660W-quad array at Centre National de Genotypage (CNG) and genotypes were called with Illumina GenomeStudio. PLINK (v1.07) was used to carry out quality control measures on an initial set of 10,015 subjects and 557,124 directly genotyped SNPs. SNPs were removed if they displayed more than 5% missingness or a Hardy-Weinberg equilibrium *p* value of less than 1.0×10^−6^. Additionally, SNPs with a minor allele frequency of less than 1% were removed. Samples were excluded if they displayed more than 5% missingness, had indeterminate X chromosome heterozygosity or extreme autosomal heterozygosity. Samples showing evidence of population stratification were identified by multidimensional scaling of genome-wide identity by state pairwise distances using the four HapMap populations as a reference, and then excluded. Cryptic relatedness was assessed using a IBD estimate of more than 0.125 which is expected to correspond to roughly 12.5% alleles shared IBD or a relatedness at the first cousin level. Related subjects that passed all other quality control thresholds were retained during subse-quent phasing and imputation. 9,048 subjects and 526,688 SNPs passed these quality control filters. The imputation was made using the IMPUTE software using the 1000 genomes phase 1 version 3 reference panel. Monomorphic or singleton SNPs were excluded. Principal components of the genetic relationship matrix were constructed using the software package GCTA (Yang et al., 2011) and using genotyped and imputed SNPs.

#### UK Biobank

Genotyping was performed using the Affymetrix UK BiLEVE Axiom array on an initial 50,000 participants; the remaining 450,000 participants were genotyped using the Affymetrix UK Biobank Axiom® array, that genotyped 850,000 variants. The two arrays are extremely sim-ilar (with over 95% common content). Quality control and imputation (to over 90 million SNPs,insertions, deletions and large structural variants) was performed by a collaborative group headed by the Wellcome Trust Centre for Human Genetics. Based on the quality control information provided, individuals were excluded based on minimal or excessive heterozygosity and disproportionate levels of individual missingness (>30%). We removed all individuals with non-European ancestry based on their own reported ethnicity together with the value of their first principal component. SNPs with a minor allele frequency of < 1%, a call rate of < 95% or evidence for violations of Hardy-Weinberg equilibrium (*p* < 1×10^−12^) were removed. Cryptic relatedness was measured as proportion of identity by descent (IBD > 0.1). Related subjects that passed all other quality control thresholds were retained during subsequent phasing and imputation. 478,840 subjects and 1,173,308 SNPs passed these quality control filters. The imputation was made using the IMPUTE software and using two different reference panels. The Haplotype Reference Consortium (HRC) panel was used wherever possible, but for SNPs not in that reference panel the UK10K + 1000 Genomes panel was used. Monomorphic or close to monomorphic (MAF < 0.001%) SNPs were excluded. Principal components of the genetic relationship matrix were provided by the UKB.

### A.3. Construction of the birth weight polygenic score

#### ALSPAC

We created polygenic scores of birth weight using GWAS summary statistics of Warrington et al. (2019). The GWAS was depleted of the ALSPAC sample, and therefore the discovery sample contained *N* = 304,057 individuals. The PGS was constructed as the sum across SNPs of the number of reference alleles for each SNP multiplied by the effect size that was corrected for linkage disequilibrium using the software package LDPred (Vilhjalmsson et al., 2015). In LDPred, a 348-kb window and a prior of 0.3 was used. In the children sample, 1,043,215 HapMap3 SNPs were used to construct the polygenic score. In the mother’s sample 1,179,707 HapMap3 SNPs were used.

#### UK Biobank

We created polygenic scores for birth weight using GWAS summary statistics of Warrington et al. (2019). The GWAS was depleted of the UK Biobank, and therefore the discovery sample contained *N* = 80,745 individuals. The PGS was constructed as the sum across SNPs of the number of reference alleles for each SNP multiplied by the effect size that was corrected for linkage disequilibrium using the software package LDPred (Vilhjalmsson et al., 2015). In LDPred, a 355-kb window and a prior of 0.3 was used. In total, 1,064,858 HapMap3 SNPs were used.

### A.4. Control variables

#### Mother’s birth weight

Mother’s self-reported their birth weight in grams. We included this measure since there is evidence that mother’s birth weight is related with their offspring birth weight (Currie and Moretti, 2007). The average birth weight of mothers is slightly lower than that of their offspring (see Table 1). For the analyses, the original continuous variable was transformed into categories (500-1,000 grams; 1,000-1,500 grams … 5,500-6,000 grams) including a category for missingness.

#### Mother’s age

Mother’s age at delivery in years. Age at delivery may be related with socio-economic status and pregnancy planning, which can be related with someone’s smoking behavior during pregnancy. On average, the mothers are 29 years old at the time of delivery. In the analyses, we included dummy variables for each age.

#### Marital status

Marital status of the mother by the time of the pregnancy. This variable is equal to 1 if the mother reported being married, and 0 in all other cases (widowed, divorced, single, separated). This measure was included since married mothers are likely to be in a relatively stable relationship with the father of the child, and therefore less subject to the stress of a possibly unwanted pregnancy. This might influence their smoking prevalence and birth outcomes. Around 80% of mothers was married.

#### Mother’s education

The education level of the mother was included. Besides being an indicator of the mother’s socio-economic status, it is also likely to be correlated with their knowledge of avail-able prenatal care. The education variable ranges from having no education (3.5% of the sample), having a CSE (certificate of secondary education) or GSE (general certificate of education, 8% of the sample), vocational training (6.2% of the sample), O-levels (36.1% of the sample), A-levels (12.5% of the sample), apprenticeship, state-enrolled or registered nurse, having a city and guilds intermediate or final technical qualification (16.8% of the sample), to having a university degree or higher (17% of the sample). An apprenticeship, being a nurse or having a city and guilds qualification were coded similarly.

#### Social class mother

This variable indicates the social class of the mother based on her occupation according to the Office of Population Censuses and Surveys (OPCS) job codes. The highest value that this variable can take is 1 (higher social class) and the lowest is 6 (lowest social class), with the aver-age in our sample being around 3. An extra category was added for armed forces which was coded as 7.

#### Grandmother’s education

The grandmother’s level of education is an indication of the family’s socioeconomic status. The education variable has categories for having CSE (certificate of secondary education), vocational training, O-levels, A-levels, or a degree. The vast majority of the maternal grandmothers had some kind of education, and the median mother had O-levels.

#### Principal components genetic relationship matrix mother

We include 4 principal components of the genetic relationship matrix to control for subtle population stratification (Price et al., 2006; Rietveld et al., 2014). Population stratification may bias the estimation of the relationship between genetic endowments and our outcome if genetic differences between subpopulations in the sample are related to unobserved factors not accounted for in the model. The inclusion of principal components is not affecting our results qualitatively, posibly because genetic outliers were already excluded from the genotyped sample and because our sample originates from a specific, relatively small, geographical area.

### A.5. OLS results (extensive margin)

#### A.5.1. ALSPAC

The OLS regressions show that, regardless of the specification, a significant relationship between maternal smoking and birth weight exists. Taking the point estimates at face value, our results suggest that self-reported maternal smoking during the first trimester is associated with a reduction in birth weight of somewhere between 146 and 174 grams. If we use cotinine levels to classify mothers into smokers and non-smokers, the association becomes weaker, with a reduction in birth weight between 56 and 99 grams.

**Table A2.** Results of the OLS regressions explaining birth weight (in ALSPAC). Coefficients are displayed with robust standard errors in parentheses. Column 1-3 use the self-reported smoking measure and Column 4-6 the self-reported smoking measure corrected using cotinine levels. All regressions correct for genetic relatedness using the first four principal components of the genetic relationship matrix.

|  | (1) | (2) | (3) | (4) | (5) | (6) |
| --- | --- | --- | --- | --- | --- | --- |
| Smoking regularly | -174.0**<br>(15.6) | -146.6**<br>(16.9) | -144.8**<br>(20.4) |  |  |  |
| Smoking regularly (adjusted) |  |  |  | -98.1**<br>(22.5) | -55.5*<br>(24.2) | -57.6*<br>(29.1) |
| Birthweight PGS (child) |  |  | 107.5**<br>(7.0) |  |  | 117.7**<br>(11.5) |
| Control variables | No | Yes | Yes | No | Yes | Yes |
| *R*-squared | 0.018 | 0.058 | 0.101 | 0.008 | 0.069 | 0.129 |
| *N* | 7,417 | 7,417 | 4,931 | 2,777 | 2,777 | 1,856 |

+ $p < 0.1$ , \* $p < 0.05$ , \*\* $p < 0.01$

#### A.5.2. UK Biobank

Table A3 presents the baseline OLS estimates in the UKB. Birth weight is measured in grams, and maternal smoking is a self-reported variable about the extensive margin of smoking (smoking mother vs. non-smoking mother). The results show a highly significant impact of both maternal smoking as well as of the PGS for birth weight on the individual’s birth weight.

**Table A3.**
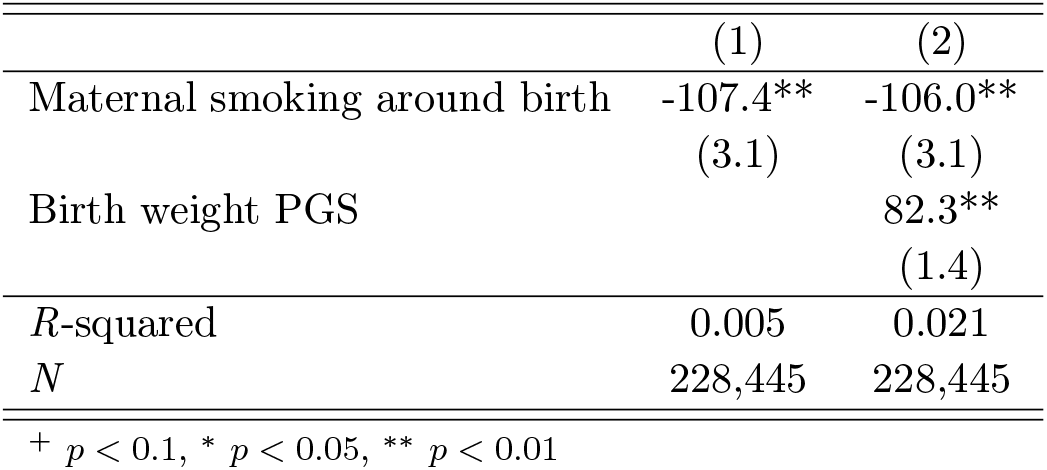
Results of the OLS regressions explaining birth weight (in the UKB). Coefficients are displayed with robust standard errors in parentheses. All regressions correct for genetic relatedness using the first 20 principal components of the genetic relationship matrix.

### A.6. Patterns in smoking reporting

In this subsection, we extend our analysis of the maternal smoking decision by assessing the characteristics of different subgroups with respect to smoking and reporting behavior using standard *t*-tests. We begin by comparing the characteristics of mothers who self-report to be smoking with those of mothers who self-report not to be smoking. Table A4 shows that mothers who report not to smoke during pregnancy, compared to mothers who report to smoke, are born slightly lighter, are about 2 and a half years older at delivery, are significantly more likely to be married, are more educated, and from a higher social. Smoking and non-smoking mother’s do not differ significantly regarding their own mother’s education.

**Table A4.** Descriptive statistics of the ALSPAC sample by self-reported smoking status.

|  | Non-smokers<br>(Self-reported) | Smokers<br>(Self-reported) | Difference |
| --- | --- | --- | --- |
| Mother's birth weight | 3,272.365<br>(597.542) | 3,218.411<br>(654.141) | -53.954**<br>(16.803) |
| Mother's age | 28.755<br>(4.691) | 26.266<br>(5.104) | -2.489**<br>(0.096) |
| Mother's marital status (Married=1) | 0.828<br>(0.377) | 0.526<br>(0.499) | -0.302**<br>(0.008) |
| Mother's education (categories) | 3.766<br>(1.593) | 2.892<br>(1.667) | -0.874**<br>(0.043) |
| Mother's social class (categories) | 2.800<br>(1.052) | 3.286<br>(1.186) | 0.486**<br>(0.026) |
| Grandmother's education (categories) | 2.438<br>(1.292) | 2.405<br>(1.290) | -0.033<br>(0.038) |
| $N$ | 9,994 | 3,365 | |

Using Table A5, we assess the characteristics of the smoking and non-smoking mothers using the self-reported smoking status corrected using our measure of cotinine. That is, mothers who report themselves to be non-smokers yet do pass the threshold of 100 ng/ml urine cotinine are now classified as smokers. Note that the sample here is restricted given that we do not have a cotinine measure for all mothers. Similarly as in Table A4, we find that mothers who smoke, compared to those who do not smoke, are born lighter, are younger at delivery, are less likely to be married, are less educated, and from a lower social class. Again, they do not differ with respect to their own mother’s education. However, the differences between the two groups are smaller than in Table A4 (with the exception of birth weight). This implies that, on average, mothers who smoke but do not report are apparently more similar to the mothers who do not smoke with respect to the characteristics in the table.

**Table A5.**
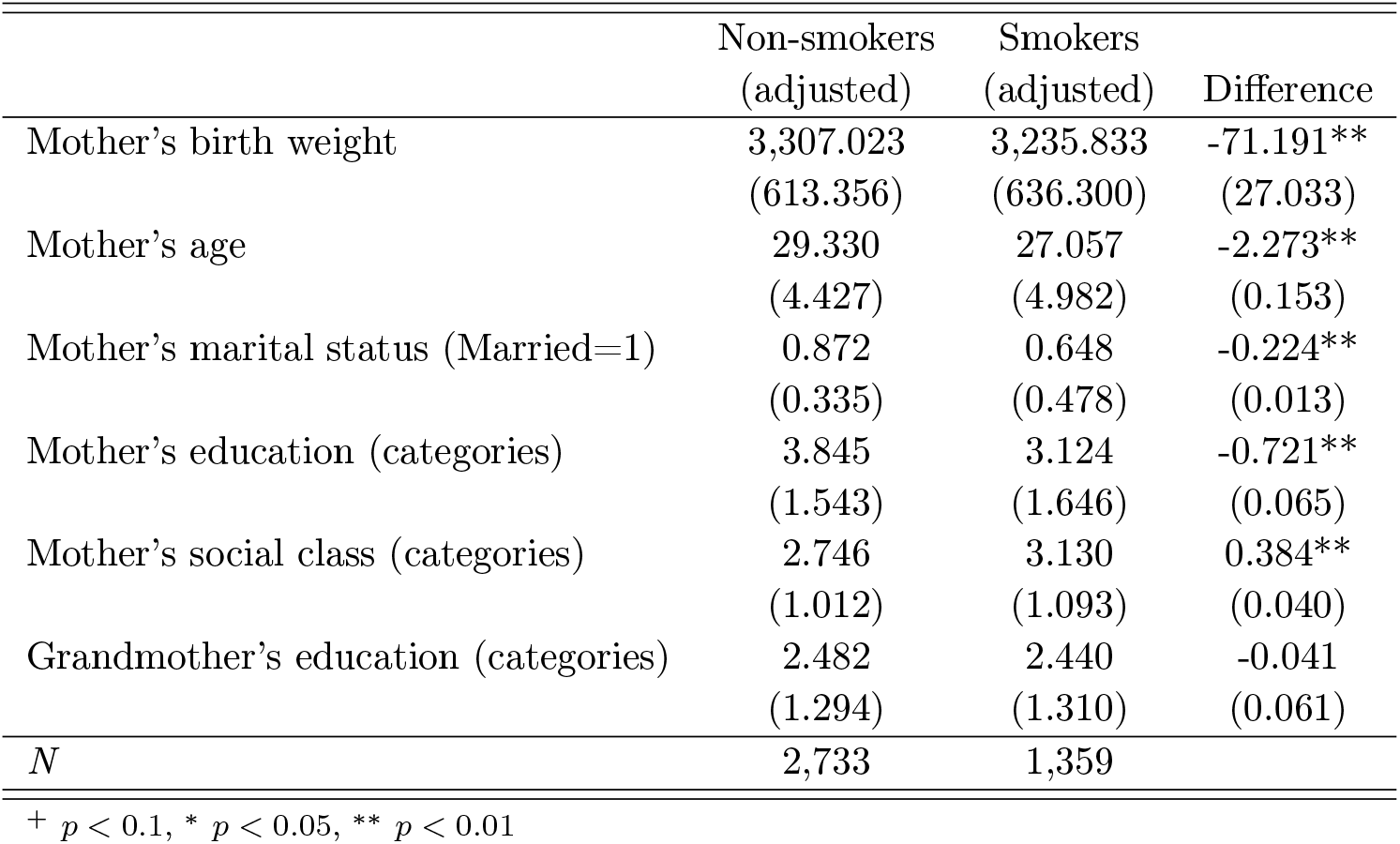
Descriptive statistics of the ALSPAC sample by self-reported smoking status adjusted using cotinine levels.

To further investigate these differences, we looked exclusively at mothers who were considered to be smokers according to our cotinine measure, and within this subsample we compared individuals who reported to smoke with the ones who reported not to smoke. Our findings are in Table A6. In this table, we also compare the number of cigarettes smoked per day (adjusted) between the two groups. We find that mothers who smoke yet do not report to smoke, compared to mothers who smoke and do report to smoke, are older at delivery, more likely to be married, more educated, from a higher social class, and their mothers are slightly higher educated. The difference in birth weight is not significant. These findings support the notion that the mothers who choose not to report even though they are actually smoking seem to be more similar to the non-smoking mothers. Interestingly, the number of cigarettes smoked adjusted on basis of the cotinine biomarker is only somewhat higher in the group who actually reports to be smoking than in the group who does not report to be smoking. These findings suggest that the measurement error in the self-reported measures of the number of cigarettes is not trivial and not easy to interpret. Therefore, we take a closer look at the first stage regression and how sensitive our results are to this source of bias in Appendix A.9.

**Table A6.** Descriptive statistics of the ALSPAC sample by self-reported smoking status. Subsample of mothers who smoke according to our cotinine measure.

|  | Non-smokers<br>(self-reported) | Smokers<br>(self-reported) | Difference |
| --- | --- | --- | --- |
| Mother's birth weight | 3,238.427<br>(608.330) | 3,234.395<br>(651.891) | -4.031<br>(48.219) |
| Mother's age | 27.540<br>(4.963) | 26.798<br>(4.975) | -0.742**<br>(0.283) |
| Mother's marital status (1=Married) | 0.771<br>(0.421) | 0.580<br>(0.494) | -0.191**<br>(0.027) |
| Mother's education (categories) | 3.434<br>(1.569) | 2.950<br>(1.664) | -0.485**<br>(0.120) |
| Mother's social class (categories) | 3.011<br>(1.032) | 3.199<br>(1.123) | 0.188*<br>(0.073) |
| Grandmother's education (categories) | 2.306<br>(1.308) | 2.522<br>(1.306) | 0.216*<br>(0.109) |
| #Cigarettes smoked per day (adjusted) | 9.044<br>(1.301) | 9.578<br>(6.483) | 0.534+<br>(0.310) |
| *N* | 474 | 885 |  |

<sup>+</sup> $p < 0.1$ , \* $p < 0.05$ , \*\* $p < 0.01$

### A.7. Independence assumption

Table A7 shows the results of the models explaining the maternal SNP rs1051730 (our instrumental variable) by each control variable separately. We report results of the *F*-test that all dummies of the categories of the control variables are equal to zero. Reassuringly, all *F*-statistics are not significantly different from zero. Hence, these results support that the independence assumption holds in our sample.

**Table A7.** Results of the OLS regressions in ALSPAC explaining the SNP rs1051730 (our instrumental variable).

|  | Mother's<br>birth weight<br>(1) | Mother's<br>age<br>(2) | Mother's<br>marital status<br>(3) | Mother's<br>education<br>(4) | Mother's<br>social class<br>(5) | Grandmother's<br>education<br>(6) |
| --- | --- | --- | --- | --- | --- | --- |
| $F$-statistic | 0.68 | 1.19 | 1.03 | 1.00 | 1.42 | 1.86 |
| $R$-squared | 0.00 | 0.00 | 0.00 | 0.00 | 0.00 | 0.00 |
| $N$ | 7,598 | 7,598 | 7,598 | 7,598 | 7,598 | 7,598 |

### A.8. Allele frequencies by smoking status

In our assessment of the exclusion restriction in section 4, we stratified the sample by smoking status and checked the relationship between our IV and the outcome in these two samples. It could however be that this endogenous stratification into smoking and non-smoking mothers biases the effect of the SNP on child’s outcomes downwards. In order to assess this issue we compare the distribution of the effect allele between smokers and non-smokers according to our cotinine measure. The results are shown in Table A8. A *t*-test for the difference in mean values between the smokers and non-smokers shows a difference of 0.035 (0.027), which is not statistically significant. Based on this result, we conclude that selection into smoking or non-smoking based on the SNP is not likely to be a major factor of concern.

**Table A8.** The distribution of A alleles of SNP rs1051370 in ALSPAC by self-reported smoking adjusted using cotinine levels.

| Number of effect alleles | Non-smokers<br>($>100\text{ng/ml}$) | Smokers<br>($<100\text{ng/ml}$) | Total |
| --- | --- | --- | --- |
| 0 | 46.3% | 42.0% | 45.0% |
| 1 | 41.7% | 46.7% | 43.2% |
| 2 | 12.1% | 11.3% | 11.8% |
| Total | 100.0% | 100.0% | 100.0% |

### A.9. Additional first stage regression results

In order to construct the adjusted number of cigarettes smoked per day, we assumed that mothers who reported to smoke a positive number of cigarettes reported their smoking behavior truthfully. In this section, we relax this assumption and assume that mothers who reported to have smoked a positive number of cigarettes also reported below the actual number they smoked. We conducted this exercise in order to evaluate how this would affect our IV estimates. Since the first stage would then increase, the overall estimate will decrease. The results are summarized in Table A9 below.

**Table A9.** Results of the sensitivity analyses relaxing the assumption of truly reporting the intensity of smoking conditional on reporting a positive number of cigarettes smoked per day. Coefficients (Coef.) are displayed with robust standard errors (S.E.) in parentheses. In the first stage regressions, all the control variables are included. The IV estimate comes from Table.

| Underreported #cigarettes | First Stage |  |  | Reduced form | IV estimate |
| --- | --- | --- | --- | --- | --- |
| | Coef. | S.E. | $p$ | | |
| 0 | 0.562** | (0.144) | 0.000 | -16.7 | -29.7 |
| 1 | 0.600** | (0.154) | 0.000 | -16.7 | -27.8 |
| 2 | 0.625** | (0.162) | 0.000 | -16.7 | -26.7 |
| 3 | 0.649** | (0.171) | 0.000 | -16.7 | -25.7 |
| 4 | 0.674** | (0.179) | 0.000 | -16.7 | -24.8 |
| 5 | 0.698** | (0.188) | 0.000 | -16.7 | -23.9 |
| 6 | 0.723** | (0.197) | 0.000 | -16.7 | -23.1 |
| 7 | 0.747** | (0.207) | 0.000 | -16.7 | -22.4 |
| 8 | 0.772** | (0.216) | 0.000 | -16.7 | -21.6 |
| 9 | 0.796** | (0.225) | 0.000 | -16.7 | -21.0 |
| 10 | 0.820** | (0.235) | 0.000 | -16.7 | -20.4 |
| $N$ | 3,852 | - | - | 7,598 | - |

<sup>+</sup> $p < 0.1$ , \* $p < 0.05$ , \*\* $p < 0.01$

The IV estimates in Table A9 are obtained by dividing the reduced form by the first stage coefficient. The main take-away is that even if we assume that all smoking mothers under-report their cigarette consumption by 10 cigarettes per day, the point estimate continues to be considerably larger than the one we obtain in the OLS regression (Table 2). It seems therefore safe to conclude that our IV point estimates exceed the OLS estimates under reasonable scenarios of measurement error in the self-reported smoking measures.

### A.10. Additional treatment effect heterogeneity analysis results

Table A10 presents the reduced form results for the ALSPAC sample that are visualized in Figure 2 in the main text.

**Table A10.** Results of the OLS (reduced form) regressions explaining birth weight for subsamples based on quartiles of the children’s PGS of birth weight. Coefficients are displayed with robust standard errors in parentheses. All regressions correct for genetic relatedness among the mothers using the first four principal components of the genetic relationship matrix

|  | Quartile of the Birth weight PGS |  |  |  |
| --- | --- | --- | --- | --- |
|  | (1) | (2) | (3) | (4) |
| rs1051370 | -10.3<br>(20.6) | 5.4<br>(21.6) | -4.3<br>(25.2) | 7.1<br>(22.4) |
| Control variables | Yes | Yes | Yes | Yes |
| *R*-squared | 0.040 | 0.045 | 0.009 | 0.037 |
| *N* | 1,252 | 1,251 | 1,252 | 1,251 |

In Table A11, we test whether the first stage was significant for the two measures of smoking. The main take away is that when we separate our sample into quartiles based on the PGS for birth weight we no longer have a strong first stage in each quartile.

**Table A11.** Results of the OLS (first stage) regressions explaining the number of cigarettes smoked per day based on quartiles of the children’s PGS of birth weight. Columns 1-4 show the results for the self reported number of cigarettes and 5-8 for the adjusted number of cigarettes. Coefficients are displayed with robust standard errors in parentheses. All regressions correct for genetic relatedness among the mothers using the first four principal components of the genetic relationship matrix.

|  | #Cigs smoked per day<br>Quartile of the<br>birth weight PGS |  |  |  | #Cigs smoked per day (adjusted)<br>Quartile of the<br>birth weight PGS |  |  |  |
| --- | --- | --- | --- | --- | --- | --- | --- | --- |
|  | (1) | (2) | (3) | (4) | (1) | (2) | (3) | (4) |
| rs1051370 | 0.8**<br>(0.2) | 0.2<br>(0.2) | 0.2<br>(0.2) | 0.3<br>(0.2) | 0.8*<br>(0.4) | 0.2<br>(0.3) | 0.1<br>(0.5) | -0.0<br>(0.4) |
| Control variables | Yes | Yes | Yes | Yes | Yes | Yes | Yes | Yes |
| *R*-squared | 0.175 | 0.161 | 0.142 | 0.190 | 0.271 | 0.260 | 0.227 | 0.310 |
| *N* | 1,252 | 1,251 | 1,252 | 1,251 | 577 | 611 | 605 | 615 |

Table A12 presents the reduced form results for the UKB sample that are visualized in Figure 2 in the main text.

**Table A12.** Results of the OLS (reduced form) regressions explaining birth weight for subsamples based on quartiles of the children’s PGS of birth weight. Coefficients are displayed with robust standard errors in parentheses. All regressions correct for genetic relatedness using the first 20 principal components of the genetic relationship matrix.

|  | Quartile of the Birth weight PGS |  |  |  |
| --- | --- | --- | --- | --- |
|  | (1) | (2) | (3) | (4) |
| rs1051370 | -0.4 | -8.8* | -10.0* | -4.5 |
|  | (3.9) | (3.9) | (3.9) | (4.0) |
| *R*-squared | 0.000 | 0.000 | 0.000 | 0.000 |
| *N* | 63,595 | 63,614 | 64,473 | 65,020 |

## Footnotes

2 See Mejdoubi et al. (2014) for an exception, where despite a modest reduction in cigarette smoking during pregnancy in an intervention group no effects on birth outcomes were found.

3 We focus in our study on the impact of genes, i.e., the sequence of molecules in the DNA which does not change over the life-time. There exists a related literature on how maternal smoking induces epigenetic expression in offspring (e.g., Joubert et al., 2016). Epigenetics is the study of gene expression (active versus inactive genes) that do not involve changes in the underlying DNA sequence. Epigenetic expression could be a mechanism through which the effect of maternal smoking arises, and additionally provides a potential mechanism through which genes and the environment interact. However, it has not been established whether epigenetic expression mediates the effect of maternal smoking on birth weight. Moreover, because of possible behavioral adjustments of the mother or child during pregnancy, epigenetic expression alone is not a sufficient condition for the presence of gene-environment interactions in birth weight.

4 There is, however, one study analyzing heterogeneity in the effect of maternal smoking on the child’s birth weight by *maternal* genotype (Wang et al., 2002), but this study focuses on maternal rather the offspring genotype, relies on specific maternal candidate genes rather than a polygenic score, and does not take into account the endogeneity of maternal smoking decisions. Moreover, Trejo (2020) analyzes interactions between the *child’s* birth weight polygenic score and maternal socio-economic status and BMI, but does not take into account the endogeneity of these maternal characteristics.

5 ALSPAC’s study website contains details of all the available data through a fully searchable data dictionary and variable search tool, see http://www.bristol.ac.uk/alspac/researchers/our-data/.

5 ALSPAC’s study website contains details of all the available data through a fully searchable data dictionary and variable search tool, see http://www.bristol.ac.uk/alspac/researchers/our-data/.

6 While nicotine has a half-life of about 2 to 3 hours – meaning that 2 hours after ingestion, half of the nicotine disappears from one’s body – cotinine has a half-life of approximately 12 to 20 hours (Kim, 2016). This implies that cotinine remains in the body for a much longer period than nicotine, which is the typical test substance in standard tobacco tests.

7 The self-reported prevalence in the subsample for which we have cotinine measurements is 20%.

8 We also included the offspring sex in our analyses as a sensitivity analysis, and results were very similar (results are available upon request from the authors).

9 Sometimes, SNP rs16969968 is used instead of SNP rs1051730. These two SNPs are in almost perfect linkage disequilibrium with each other, meaning that they are almost perfectly correlated with each other in the human genome.

10 Of course, there is a mechanical correlation between the maternal SNP and the child’s polygenic score since the child’s birth weight polygenic score includes all SNPs, including rs1051730. However, this SNP only represents one out of many SNPs in the polygenic score and therefore the correlation is negligible. Moreover, we performed a robustness check in which we excluded the entire genetic region in which SNP rs1051730 resides from the polygenic score and this did not alter the results in any way.

11 That is, since the SNP is related to nicotine dependence, it may also determine subgroup membership (smoker or non-smoker). If true, this would bias the effect among non-smokers towards zero, since this is a group of resilient mothers who despite having the SNP do not smoke. However, the effect of the SNP on the binary indicator of smoking is much smaller than its effect on smoking intensity, consistent with the fact that around 80% of smokers start before they are 18 years old, and that smoking initiation is influenced mainly by environmental factors rather than genes.

12 Ideally, we would have observed cotinine levels for all 7,598 mothers throughout the entire pregnancy, but we are limited to 2,844 measurements from the first trimester only (see Table 1). Therefore, we use the cotinine measure only to gauge the bias in the first stage, and use baseline sample 1 whenever possible to maximize the sample size.

13 In the reduced form, therefore, we effectively interact the maternal SNP with the child’s PGS which constitutes a so-called Gene-Gene (*G* × *G*) interaction. However, since we provide evidence below that plausibly the single channel through which the maternal SNP impacts the child’s outcomes is maternal smoking during pregnancy, this *G* × *G* interaction effectively reflects a Gene-Environment (*G* × *E*) interaction.

14 A similar decrease is still apparent when we analyze the self-reported smoking measure in the reduced sample for which we observe cotinine measurements (results are available upon request from the authors).

15 An alternative explanation could be that since our cotinine variable comes from a single measurement somewhere during pregnancy, it is a poorer measure of the average number of cigarettes per day smoked throughout pregnancy than the self-reported measure. However, this does not explain the pattern for the extensive margin of smoking in Appendix A.5 since in this case updating the smoking status with this measure implies an unambiguous improvement with respect to the original classification.

16 Stratifying on basis of the self-reported smoking status gives very similar results, which are available upon request from the authors.

17 This is because in the IV estimation, only the first stage includes the number of cigarettes smoked, and the resulting attenuated coefficient appears in the denominator of the 2SLS estimates and thus inflates the IV estimate.

18 Since the UKB relies on voluntary participation, there is a small subsample of mother-child pairs who both volunteered to participate. This subsample consists of approximately 4,400 families (Young et al., 2020). This subsample is thus smaller than our main analysis sample from ALSPAC, and therefore it is not suitable to assess the robustness of our results.

19 Our results are consistent with those of Yang et al. (2019), and very close in magnitude to what they have found:-18 grams for individuals with a smoking mother and −2 grams for individuals with a non-smoking mother. The small differences might be explained by the fact that the authors used the SNP rs16969968 as a proxy instrument. This SNP is in almost perfect linkage disequilibrium (LD) with rs1051730.

## References

Aizer, A. (2011). Poverty, violence, and health the impact of domestic violence during pregnancy on newborn health. Journal of Human Resources, 46(3):518–538.

Almond, D., Chay, K. Y., and Lee, D. S. (2005). The costs of low birth weight. The Quarterly Journal of Economics, 120(3):1031–1083.

Almond, D. and Currie, J. (2011). Killing me softly: The fetal origins hypothesis. Journal of Economic Perspectives, 25(3):153–72.

Almond, D., Currie, J., and Duque, V. (2018). Childhood circumstances and adult outcomes: Act ii. Journal of Economic Literature, 56(4):1360–1446.

Almond, D., Hoynes, H. W., and Schanzenbach, D. W. (2011). Inside the war on poverty: The impact of food stamps on birth outcomes. The Review of Economics and Statistics, 93(2):387–403.

Andrews, I., Stock, J. H., and Sun, L. (2019). Weak instruments in instrumental variables regression: Theory and practice. Annual Review of Economics, 11:727–753.

Banderali, G., Martelli, A., Landi, M., Moretti, F., Betti, F., Radaelli, G., Lassandro, C., and Verduci, E. (2015). Short and long term health effects of parental tobacco smoking during pregnancy and lactation: a descriptive review. Journal of Translational Medicine, 13(1):1–7.

Barber, S. L. and Gertler, P. J. (2008). The impact of mexico’s conditional cash transfer programme, oportunidades, on birthweight. Tropical Medicine & International Health, 13(11):1405–1414.

Barcellos, S. H., Carvalho, L. S., and Turley, P. (2018). Education can reduce health differences related to genetic risk of obesity. Proceedings of the National Academy of Sciences, 115(42):E9765–E9772.

Beauchamp, J. P., Cesarini, D., Johannesson, M., van der Loos, M. J., Koellinger, P. D., Groenen, P. J., Fowler, J. H., Rosenquist, J. N., Thurik, A. R., and Christakis, N. A. (2011). Molecular genetics and economics. Journal of Economic Perspectives, 25(4):57–82.

Benowitz, N. L., Bernert, J. T., Caraballo, R. S., Holiday, D. B., and Wang, J. (2009). Optimal serum cotinine levels for distinguishing cigarette smokers and nonsmokers within different racial/ethnic groups in the united states between 1999 and 2004. American Journal of Epidemiology, 169(2):236– 248.

Bharadwaj, P., Johnsen, J. V., and Løken, K. V. (2014). Smoking bans, maternal smoking and birth outcomes. Journal of Public Economics, 115:72–93.

Bharadwaj, P., Lundborg, P., and Rooth, D.-O. (2018). Birth weight in the long run. Journal of Human Resources, 53(1):189–231.

Bierut, L. and Cesarini, D. (2015). How genetic and other biological factors interact with smoking decisions. Big data, 3(3):198–202.

Bierut, L. J. (2010). Convergence of genetic findings for nicotine dependence and smoking related diseases with chromosome 15q24-25. Trends in Pharmacological Sciences, 31(1):46–51.

Black, S. E., Devereux, P. J., and Salvanes, K. G. (2007). From the cradle to the labor market? the effect of birth weight on adult outcomes. The Quarterly Journal of Economics, 122(1):409–439.

Black, S. E., Devereux, P. J., and Salvanes, K. G. (2016). Does grief transfer across generations? bereavements during pregnancy and child outcomes. American Economic Journal: Applied Eco- nomics, 8(1):193–223.

Boyd, A., Golding, J., Macleod, J., Lawlor, D. A., Fraser, A., Henderson, J., Molloy, L., Ness, A. R., Ring, S. M., and Davey Smith, G. (2013). Cohort Profile: The ‘Children of the 90s’–the index offspring of the Avon Longitudinal Study of Parents and Children. International Journal of Epidemiology, 42(1):111–127.

Bradford, W. D. (2003). Pregnancy and the demand for cigarettes. American Economic Review, 93(5):1752–1763.

Bycroft, C., Freeman, C., Petkova, D., Band, G., Elliott, L. T., Sharp, K., Motyer, A., Vukcevic, D., Delaneau, O., O’Connell, J., et al. (2017). Genome-wide genetic data oñ 500,000 uk biobank participants. BioRxiv, page 166298.

Chabris, C. F., Lee, J. J., Cesarini, D., Benjamin, D. J., and Laibson, D. I. (2015). The Fourth Law of Behavior Genetics. Current Directions in Psychological Science, 24(4):304–312.

Conti, G., Hanson, M., Inskip, H., Crozier, S., Cooper, C., and Godfrey, K. M. (2018). Beyond birthweight: The origins of human capital. IZA Discussion Paper, 13296.

Currie, J. (2009). Healthy, wealthy, and wise: Socioeconomic status, poor health in childhood, and human capital development. Journal of Economic Literature, 47(1):87–122.

Currie, J. and Moretti, E. (2007). Biology as destiny? short-and long-run determinants of intergen- erational transmission of birth weight. Journal of Labor Economics, 25(2):231–264.

Currie, J. and Rossin-Slater, M. (2013). Weathering the storm: Hurricanes and birth outcomes. Journal of Health Economics, 32(3):487–503.

Davey Smith, G. (2003). ‘Mendelian randomization’: can genetic epidemiology contribute to under- standing environmental determinants of disease? International Journal of Epidemiology, 32(1):1–22.

Davey Smith, G., Lawlor, D. A., Harbord, R. M., Timpson, N. J., Day, I. N. M., and Ebrahim, S. (2007). Clustered environments and randomized genes: a fundamental distinction between conven- tional and genetic epidemiology. PLoS medicine, 4(12):e352.

de Weerd, S., Thomas, C. M., Kuster, J. E., Cikot, R. J., and Steegers, E. A. (2002). Variation of serum and urine cotinine in passive and active smokers and applicability in preconceptional smoking cessation counseling. Environmental Research, 90(2):119–124.

Doyle, M.-A., Schurer, S., and Silburn, S. (2020). Unintended consequences of welfare reform: Evi- dence from birth outcomes of aboriginal australians.

Dudbridge, F. (2013). Power and predictive accuracy of polygenic risk scores. PLoS Genetics, 9(3):e1003348.

England, L. J., Kendrick, J. S., Gargiullo, P. M., Zahniser, S. C., and Hannon, W. H. (2001). Measures of maternal tobacco exposure and infant birth weight at term. American Journal of Epidemiology, 153(10):954–960.

Evans, W. N. and Ringel, J. S. (1999). Can higher cigarette taxes improve birth outcomes? Journal of Public Economics, 72(1):135–154.

Figlio, D., Guryan, J., Karbownik, K., and Roth, J. (2014). The effects of poor neonatal health on children’s cognitive development. American Economic Review, 104(12):3921–55.

Fraser, A., Macdonald-Wallis, C., Tilling, K., Boyd, A., Golding, J., Davey Smith, G., Henderson, J., Macleod, J., Molloy, L., Ness, A., et al. (2013). Cohort profile: the avon longitudinal study of parents and children: Alspac mothers cohort. International Journal of Epidemiology, 42(1):97–110.

Furberg, H., Kim, Y., Dackor, J., Boerwinkle, E., Franceschini, N., Ardissino, D., Bernardinelli, L., Mannucci, P. M., Mauri, F., Merlini, P. A., et al. (2010). Genome-wide meta-analyses identify multiple loci associated with smoking behavior. Nature Genetics, 42(5):441.

Hamilton, B. H. (2001). Estimating treatment effects in randomized clinical trials with non- compliance: the impact of maternal smoking on birthweight. Health Economics, 10(5):399–410.

Heckman, J. J. (2007). The economics, technology, and neuroscience of human capability formation. Proceedings of the National Academy of Sciences, 104(33):13250–5.

Hoffmann, D., Haley, N. J., Adams, J. D., and Brunnemann, K. D. (1984). Tobacco sidestream smoke: uptake by nonsmokers. Preventive Medicine, 13(6):608–617.

Horikoshi, M., Beaumont, R. N., Day, F. R., Warrington, N. M., …, and Freathy, R. M. (2016). Genome-wide associations for birth weight and correlations with adult disease. Nature, 538(7624):248–252.

Horikoshi, M., Yaghootkar, H., Mook-Kanamori, D. O., Sovio, U., Taal, H. R., Hennig, B. J., Bradfield, J. P., St Pourcain, B., Evans, D. M., Charoen, P., et al. (2013). New loci associated with birth weight identify genetic links between intrauterine growth and adult height and metabolism. Nature Genetics, 45(1):76–82.

Joubert, B. R., Felix, J. F., Yousefi, P., Bakulski, K. M., Just, A. C., Breton, C., Reese, S. E., Markunas, C. A., Richmond, R. C., Xu, C.-J., et al. (2016). Dna methylation in newborns and maternal smoking in pregnancy: genome-wide consortium meta-analysis. The American Journal of Human Genetics, 98(4):680–696.

Kataoka, M. C., Carvalheira, A. P. P., Ferrari, A. P., Malta, M. B., Carvalhaes, M. A. d. B. L., and de Lima Parada, C. M. G. (2018). Smoking during pregnancy and harm reduction in birth weight: a cross-sectional study. BMC Pregnancy and Childbirth, 18(1):1–10.

Kidd, J. M., Cooper, G. M., Donahue, W. F., Hayden, H. S., Sampas, N., Graves, T., Hansen, N., Teague, B., Alkan, C., Antonacci, F., et al. (2008). Mapping and sequencing of structural variation from eight human genomes. Nature, 453(7191):56–64.

Kim, S. (2016). Overview of cotinine cutoff values for smoking status classification. International Journal of Environmental Research and Public Health, 13(12):1236.

Kong, A., Thorleifsson, G., Frigge, M. L., Vilhjalmsson, B. J., Young, A. I., Thorgeirsson, T. E., Benonisdottir, S., Oddsson, A., Halldorsson, B. V., Masson, G., et al. (2018). The nature of nurture: Effects of parental genotypes. Science, 359(6374):424–428.

Kramer, M. S. (1987). Intrauterine growth and gestational duration determinants. Pediatrics, 80(4):502–511.

Langone, J. J., Gjika, H. B., and Van Vunakis, H. (1973). Nicotine and its metabolites. radioim- munoassays for nicotine and cotinine. Biochemistry, 12(24):5025–5030.

Lawlor, D., Richmond, R., Warrington, N., McMahon, G., Smith, G. D., Bowden, J., and Evans, D. M. (2017). Using mendelian randomization to determine causal effects of maternal pregnancy (intrauterine) exposures on offspring outcomes: Sources of bias and methods for assessing them. Wellcome Open Research, 2.

Lehrer, S. F. and Ding, W. (2017). Are genetic markers of interest for economic research? IZA Journal of Labor Policy, 6(1):2.

Li, C. Q., Windsor, R. A., Perkins, L., Goldenberg, R. L., and Lowe, J. B. (1993). The impact on infant birth weight and gestational age of cotinine-validated smoking reduction during pregnancy. JAMA, 269(12):1519–1524.

Lien, D. S. and Evans, W. N. (2005). Estimating the impact of large cigarette tax hikes the case of maternal smoking and infant birth weight. Journal of Human Resources, 40(2):373–392.

Lindqvist, R., Lendahls, L., Tollbom, Ö., åBerg, H., and Håkansson, A. (2002). Smoking during preg- nancy: comparison of self-reports and cotinine levels in 496 women. Acta Obstetricia et Gynecologica Scandinavica, 81(3):240–244.

Liu, J. Z., Tozzi, F., Waterworth, D. M., Pillai, S. G., Muglia, P., Middleton, L., Berrettini, W., Knouff, C. W., Yuan, X., Waeber, G., et al. (2010). Meta-analysis and imputation refines the association of 15q25 with smoking quantity. Nature Genetics, 42(5):436–440.

Liu, M., Jiang, Y., Wedow, R., Li, Y., Brazel, D. M., Chen, F., Datta, G., Davila-Velderrain, J., McGuire, D., Tian, C., et al. (2019). Association studies of up to 1.2 million individuals yield new insights into the genetic etiology of tobacco and alcohol use. Nature Genetics, 51(2):237–244.

Mejdoubi, J., van den Heijkant, S. C., van Leerdam, F. J., Crone, M., Crijnen, A., and HiraSing, R. A. (2014). Effects of nurse home visitation on cigarette smoking, pregnancy outcomes and breastfeeding: a randomized controlled trial. Midwifery, 30(6):688–695.

Mills, M. C., Barban, N., and Tropf, F. C. (2020). An Introduction to Statistical Genetic Data Analysis. MIT Press.

Olea, J. L. M. and Pflueger, C. (2013). A robust test for weak instruments. Journal of Business & Economic Statistics, 31(3):358–369.

Oyesanya, O. A., Zenzes, M. T., Wang, P., and Casper, R. F. (1995). Cigarette smoking may affect meiotic maturation of human oocytes. Human Reproduction, 10(12):3213–3217.

Persson, P. and Rossin-Slater, M. (2018). Family ruptures, stress, and the mental health of the next generation. American Economic Review, 108(4-5):1214–52.

Polderman, T. J., Benyamin, B., De Leeuw, C. A., Sullivan, P. F., Van Bochoven, A., Visscher, P. M., and Posthuma, D. (2015). Meta-analysis of the heritability of human traits based on fifty years of twin studies. Nature Genetics, 47(7):702–709.

Price, A. L., Patterson, N. J., Plenge, R. M., Weinblatt, M. E., Shadick, N. A., and Reich, D. (2006). Principal components analysis corrects for stratification in genome-wide association studies. Nature Genetics, 38(8):904–909.

Rietveld, C. A., Conley, D., Eriksson, N., Esko, T., Medland, S. E., Vinkhuyzen, A. A., Yang, J., Boardman, J. D., Chabris, C. F., Dawes, C. T., et al. (2014). Replicability and robustness of genome-wide-association studies for behavioral traits. Psychological Science, 25(11):1975–1986.

Ringel, J. S. and Evans, W. N. (2001). Cigarette taxes and smoking during pregnancy. American Journal of Public Health, 91(11):1851–1856.

Royer, H. (2009). Separated at girth: Us twin estimates of the effects of birth weight. American Economic Journal: Applied Economics, 1(1):49–85.

Rutter, M. (2006). Genes and Behavior: Nature-Nurture Interplay Explained. Blackwell Publishers, Oxford, UK.

Schmitz, L. L. and Conley, D. (2017). The effect of vietnam-era conscription and genetic potential for educational attainment on schooling outcomes. Economics of Education Review, 61:85–97.

Sexton, M. and Hebel, J. R. (1984). A clinical trial of change in maternal smoking and its effect on birth weight. JAMA, 251(7):911–915.

Simon, D. (2016). Does early life exposure to cigarette smoke permanently harm childhood welfare? evidence from cigarette tax hikes. American Economic Journal: Applied Economics, 8(4):128–59.

Tappin, D., Bauld, L., Purves, D., Boyd, K., Sinclair, L., MacAskill, S., McKell, J., Friel, B., Mc- Connachie, A., De Caestecker, L., et al. (2015). Financial incentives for smoking cessation in pregnancy: randomised controlled trial. British Medical Journal, 350.

Tappin, D., Ford, R., and Schluter, P. (1997). Smoking during pregnancy measured by population cotinine testing. The New Zealand Medical Journal, 110(1050):311–314.

The 1000 Genomes Project Consortium (2015). A global reference for human genetic variation. Nature, 526(7571):68–74.

Trejo, S. (2020). Exploring genetic influences on birth weight. SocArXiv, 10.31235/osf.io/7j59q.

Turkheimer, E. (2000). Three laws of behavior genetics and what they mean. Current Directions in Psychological Science, 9(5):160–164.

Tyrrell, J., Huikari, V., Christie, J. T., Cavadino, A., Bakker, R., Brion, M.-J. A., Geller, F., Pater-noster, L., Myhre, R., Potter, C., et al. (2012). Genetic variation in the 15q25 nicotinic acetylcholine receptor gene cluster (chrna5–chrna3–chrnb4) interacts with maternal self-reported smoking status during pregnancy to influence birth weight. Human Molecular Genetics, 21(24):5344–5358.

Van Kippersluis, H. and Rietveld, C. A. (2018a). Beyond plausibly exogenous. The Econometrics Journal, 21(3):316–331.

Van Kippersluis, H. and Rietveld, C. A. (2018b). Pleiotropy-robust mendelian randomization. Inter- national Journal of Epidemiology, 47(4):1279–1288.

Vilhjálmsson, B. J., Yang, J., Finucane, H. K., Gusev, A., Lindström, S., Ripke, S., Genovese, G., Loh, P.-R., Bhatia, G., Do, R., et al. (2015). Modeling linkage disequilibrium increases accuracy of polygenic risk scores. The American Journal of Human Genetics, 97(4):576–592.

Vilhjalmsson, B. J., Yang, J., Finucane, H. K., Gusev, A., Lindström, S., Ripke, S., Genovese, G., Loh, P.-R., Bhatia, G., Do, R., Hayeck, T., Won, H.-H., Kathiresan, S., Pato, M., Pato, C., Tamimi, R., Stahl, E. A., Zaitlen, N., Pasaniuc, B., Belbin, G., Kenny, E. E., Schierup, M. H., De Jager, P. L., Patsopoulos, N. A., McCarroll, S. A., Daly, M. J., Purcell, S. M., Chasman, D. I., Neale, B. M., Goddard, M., Visscher, P. M., Kraft, P., Patterson, N., and Price, A. L. (2015). Modeling Linkage Disequilibrium Increases Accuracy of Polygenic Risk Scores. The American Journal of Human Genetics, 97(4):576–592.

Visscher, P. M., Wray, N. R., Zhang, Q., Sklar, P., McCarthy, M. I., Brown, M. A., and Yang, J. (2017). 10 Years of GWAS Discovery: Biology, Function, and Translation. The American Journal of Human Genetics, 101(1):5–22.

von Hinke, S., Smith, G. D., Lawlor, D. A., Propper, C., and Windmeijer, F. (2016). Genetic markers as instrumental variables. Journal of Health Economics, 45:131–148.

Wang, X., Tager, I. B., Van Vunakis, H., Speizer, F. E., and Hanrahan, J. P. (1997). Maternal smoking during pregnancy, urine cotinine concentrations, and birth outcomes. a prospective cohort study. International Journal of Epidemiology, 26(5):978–988.

Wang, X., Zuckerman, B., Pearson, C., Kaufman, G., Chen, C., Wang, G., Niu, T., Wise, P. H., Bauchner, H., and Xu, X. (2002). Maternal cigarette smoking, metabolic gene polymorphism, and infant birth weight. JAMA, 287(2):195–202.

Warrington, N. M., Beaumont, R. N., Horikoshi, M., Day, F. R., Helgeland, Ø., Laurin, C., Bacelis, J., Peng, S., Hao, K., Feenstra, B., et al. (2019). Maternal and fetal genetic effects on birth weight and their relevance to cardio-metabolic risk factors. Nature Genetics, 51(5):804–814.

Wehby, G. L., Fletcher, J. M., Lehrer, S. F., Moreno, L., Murray, J. C., Wilcox, A., and Lie, R. (2011). A Genetic Instrumental Variables Analysis of the Effects of Prenatal Smoking on Birth Weight: Evidence from Two Samples. Biodemography and social biology, 57(1):3–32.

Yang, J., Lee, S. H., Goddard, M. E., and Visscher, P. M. (2011). Gcta: a tool for genome-wide complex trait analysis. The American Journal of Human Genetics, 88(1):76–82.

Yang, Q., Millard, L. A., and Davey Smith, G. (2019). Proxy gene-by-environment mendelian ran- domization study confirms a causal effect of maternal smoking on offspring birthweight, but little evidence of long-term influences on offspring health. International Journal of Epidemiology.

Young, A. I., Nehzati, S. M., Lee, C., Benonisdottir, S., Cesarini, D., Benjamin, D. J., Turley, P., and Kong, A. (2020). Mendelian imputation of parental genotypes for genome-wide estimation of direct and indirect genetic effects. BioRxiv.

Zheng, W., Suzuki, K., Tanaka, T., Kohama, M., Yamagata, Z., and Group, O. C. H. S. (2016). Association between maternal smoking during pregnancy and low birthweight: effects by maternal age. PLOS ONE, 11(1):e0146241.

